# Genetic Profiling of Autoimmune Diseases and Exploring Clusters Through Polygenic Risk Score Analysis Using Cohort Data from the UK Biobank

**DOI:** 10.64898/2026.05.09.26352677

**Authors:** Rochi Saurabh, Inken Wohlers, Marius Möller, Hauke Busch

## Abstract

Autoimmune diseases result from immune responses against self-antigens but exhibit marked phenotypic diversity shaped by genetic and environmental factors. Genome-wide association studies (GWAS) have identified susceptibility loci that inform polygenic scores (PGS) for risk prediction. This study integrates phenotypic and genetic data from the UK Biobank(UKB) to characterize disease overlap, genetic heterogeneity, and shared biological mechanisms across autoimmune conditions. Comorbidity patterns were further assessed using patient records from UKB and the TriNetX(TNX). Phenotypic data from 502,371 UKB participants were used to evaluate diagnostic overlap, with a subset of 104,544 individuals analyzed for PGS distributions. Significant variants were identified using genome-wide thresholds, allele frequency, and predicted impact, and shared genes were subsequently mapped to pathways using Hallmark gene sets. Comorbidity across rare and common autoimmune diseases was assessed in the UKB and TNX using ICD-10 codes, focusing on White individuals (71,069,654 in TNX; 502,371 in UKB). Odds ratios for 15 diseases were estimated, and cross-cohort comparisons evaluated reproducibility and cohort-specific differences. PGS analyses revealed both shared and distinct genetic architectures, indicating partial genetic overlap and supporting poly-autoimmunity. Integration of common, rare and impactful variants identified both known and novel gene associations, while pathway analysis highlighted systemic and tissue-specific immune dysregulation. Cross-dataset comparisons confirmed consistent comorbidity patterns but underscored the impact of dataset-specific factors, emphasizing the need for standardized approaches in autoimmune disease research.

## 1 Epidemiology of Autoimmune Diseases

Immune dysregulation leads to the destruction of self-tissues in various chronic conditions known as autoimmune diseases (ADs). The abnormal activation of the immune system causes the detection of self-antigens as foreign, leading to chronic inflammation and tissue damage. Researchers have identified a common mechanism loss of self-tolerance that is fundamental to various ADs [1]. Despite clinical variability, many ADs exhibit similar pathogenetic characteristics, suggesting shared genetic and environmental influences. Furthermore, epigenetic changes and hormonal factors add to the variety and phenotypic variability seen in these disorders. According to Ngo et. al., [2], women are more predisposed than men to ADs, a difference potentially influenced by sex hormones, androgens inhibit autoimmunity while estrogens enhance it [3].

These diseases affect about 10% of people worldwide, and their incidence is on the rise, particularly for conditions like type 1 diabetes (T1D) [4]. Countries with advanced economies are seeing a rise in cases of ADs, particularly among middle-aged adults aged 40 to 50 [5, 6]. The overall prevalence of ADs is 4.5%, affecting 6.4% of females and 2.7% of males [6]. Systemic lupus erythematosus (SLE) affects women about nine times more than men, rheumatoid arthritis (RA) and multiple sclerosis (MS) affect women about three times more, and inflammatory Bowel Disease (IBD) and T1D affect women equally compared to men [7]. Around 80 unique autoimmune disorders have been recognized [8], some affecting specific organs and others presenting more generally.

Recent research highlights several additional epidemiological features that are essential for a comprehensive understanding of AD. Notably, the prevalence of multiple ADs in a single individual is substantial in a large U.S. cohort from 2011–2022, 34% of those diagnosed with an AD had more than one such diagnosis, with 24% experiencing two ADs, 8% three, and 2% four or more [9]. This phenomenon referred to as polyautoimmunity and, when three or more conditions are present, as multiple autoimmune syndrome (MAS) indicates both significant comorbidity and the need for continued surveillance, as patients with one autoimmune disorder are at higher risk for developing others [10]. Moreover, recent global data signal an alarming rise in the incidence and prevalence of ADs. Studies document a yearly worldwide incidence increase of 19.1% and a 12.5% annual rise in prevalence, affecting diverse populations and age groups [11].

Several case studies have reported that individuals with one AD are at an increased risk of developing additional autoimmune conditions [12]. Thus, genetic predispositions that lead to ADs can be discussed together. Diagnosing ADs can be challenging due to their overlapping symptoms with other conditions. Additionally, the wide range of clinical manifestations and lack of specific diagnostic tests make it difficult for healthcare professionals to pinpoint the exact autoimmune disorder. This often leads to delays in diagnosis and treatment, impacting patient outcomes.

### 1.1 Modern Genetic Approaches to Autoimmune Disease Susceptibility

Autoimmunity is strongly influenced by genetic variation. Genomic variations differences in DNA sequences across a genome can markedly affect disease susceptibility and treatment response [13]. Due to disease heterogeneity, multiple variants contribute to AD risk [14]. While common variants generally have mild effects, rare variants, particularly in coding regions, may have larger impacts and account for part of the missing heritability [15, 16]. However, their contribution remains poorly understood, highlighting the need for deeper investigation to improve genetic risk prediction. More than 90% of AD associated variants lie in non-coding regions, complicating functional interpretation [17]. Recent studies reveal shared loci across ADs, suggesting common mechanisms underlying immune tolerance breakdown [18, 19]. Despite extensive progress, linking genetic findings to disease mechanisms and clinical application remains a major challenge [20].

Genome-wide association studies (GWAS) identify genetic variations linked to diseases by comparing allele frequencies or genotypes between cases and controls [20]. Typically, hundreds of thousands of single-nucleotide polymorphisms (SNPs) are genotyped in large cohorts [21]. Although primarily focused on SNPs, GWAS have uncovered numerous disease-associated loci [22], deepening understanding of genetic contributions to human disease [23]. Over the past two decades since the first GWAS was conducted [24], more than 5,700 studies have identified genetic variants associated with over 3,300 traits [21]. However, identifying causal variants and interpreting results remain challenging [25]. In ADs, GWAS have revealed shared and disease-specific loci such as the HLA region and PTPN22 impairing immune tolerance across cell types [26]. Despite limited predictive power for individual variants, combining genetic profiles with clinical factors improves risk assessment [27].

Beyond GWAS, whole exome sequencing (WES) and whole genome sequencing (WGS) provide broader insights. WES captures over 95% of exons, encompassing 85% of known disease-causing mutations, at lower cost than WGS [28, 29]. WGS, covering all genomic variants, has enabled comprehensive global reference datasets [30]. These approaches increased the number of AD associated loci from 15 to 68 [31] and identified both coding and noncoding variants [32]. While the contribution of rare variants remains unclear [33], integrating WGS and WES findings enhances understanding of AD pathogenesis and supports translation of genetic discoveries into clinical applications.

The identification of genetic variants associated with ADs through GWAS and sequencing studies provides valuable insights into disease mechanisms. However, understanding how these variants collectively contribute to an individual’s overall disease risk requires integrative approaches. Polygenic Scores (PGS) quantify an individual’s genetic predisposition to disease, reflecting the polygenic architecture of complex traits where numerous variants each exert small effects [34, 35]. Using GWAS summary statistics, PGS aggregates the weighted effects of thousands of SNPs to estimate overall genetic risk [22, 36]. Each variant’s weight corresponds to its GWAS-derived effect size, and large-scale studies identify the variants contributing to disease susceptibility [37]. PGS has been applied across ADs including SLE, RA, IBD, T1D, vitiligo, and Sjögren’s syndrome [38]. It outperforms traditional biomarkers for risk prediction and supports early detection and treatment optimization [39, 40, 41]. High PGS scores correlate with earlier onset, severe outcomes, and organ damage in SLE [39, 42]. Integrating PGS with clinical, biochemical, and demographic factors enhances predictive performance [43, 44]. However, most PGS models are derived from European ancestry cohorts, limiting generalization to diverse populations and raising ethical and regulatory considerations for clinical application [45, 46]. Despite major advances in AD research through GWAS and PGS, key knowledge gaps persist. Many studies fail to apply a comprehensive framework that systematically ranks genetic variants by integrating allele frequency with functional annotations from tools such as the variant effect predictor (VEP). This limitation hampers understanding of AD genetics, especially the roles of both common and rare variants [47]. Although many associated variants have been identified, most studies focus on these variants in isolation rather than linking them to biological processes. Lacking this context restricts insight into disease mechanisms, and efforts to identify shared pathways across autoimmune disorders remain limited.

### 1.2 Genetic, Phenotypic Information and Autoimmune Diseases Covered by the UK Biobank and TriNetX

Between 2006 to 2010, over 500,000 individuals from the United Kingdom, aged 40 to 69 years, were registered in the UK Biobank (UKB), establishing the one of the largest collections of genetic and phenotypic data for global academic health research [30]. There are numerous databases on autoimmune illnesses, making it an effective tool for learning about their genetic foundation and clinical characteristics. The UKB has a range of phenotypic data, including height, weight, BMI, physical fitness tests, dietary habits, smoking and drinking, as well as self-reported lifestyle, medical history, and environmental exposures. For all participants, genome-wide genotyping was done using the UKB Axiom Array, which directly measured about 850,000 variants. The Haplotype Reference Consortium and UK10K + 1000 Genomes reference panels were used to fill in over 90 million missing variants. WES has been conducted on more than 470,000 participants, identifying roughly 12 million protein-coding variants. A 2021 dataset includes exome data from 454,787 individuals, comprising about 2 million exonic SNVs [48]. By 2023, sequencing had been completed for 500,000 genomes, following the release of 200,000 genomes in 2021 [38]. Sequencing was performed using Illumina NovaSeq 6000 machines with S4 flowcells for paired end reads. GWAS analyses incorporated both array-based and imputed genomic data (fields 22418 and 21008, respectively) [38]. The UKB offers a robust platform for the development of PGS and the advancement of precision medicine by amalgamating genetic, phenotypic, and environmental data [49].

TriNetX (TNX) is a real-time, federated health research network that aggregates de-identified electronic health record (EHR) data from hospitals, clinics, and research institutions worldwide. Unlike traditional biobanks, TNX does not collect data directly from participants; instead, it harmonizes EHR-derived information demographics, diagnoses, procedures, medications, laboratory results, oncology data, and genetic variants within a standardized common data model [50]. Data integration relies on custom connectors and natural language processing to extract both structured and unstructured clinical information. The federated architecture keeps data within each healthcare organization’s secure environment while enabling compliant queries under HIPAA and GDPR standards. Through continuous updates and rigorous quality control, TNX maintains missing data rates below 5% and ensures validity via a certified de-identification process [51, 50]. As of 2022, the network included over 220 healthcare organizations across 30 countries, facilitating more than 19,000 sponsored clinical trial opportunities [52, 50]. By supporting real-world evidence research, cohort identification, and data-driven trial design, the platform enhances clinical discovery and collaboration among research and industry partners.

Building on current knowledge of genetic susceptibility, polygenic risk modeling, and comprehensive AD datasets, this study aims to investigate (1) Examination of paired AD risk scores in the UKB enables the identification of clustering patterns, shared genetic variants, and common genes involved in biological pathways. (2) and explore patterns of AD comorbidities using the extensive datasets provided by UKB and TNX.

## 2 Materials and Methods

### 2.1 Analytical Framework for Integrating Phenotypic Data, Risk Scores Correlations and Profiling Across Datasets

This analysis categorizes and analyzes autoimmune disease (AD) samples using phenotype data from the initial UK Biobank (UKB) release, obtained on March 18, 2024. The included ADs align with those analyzed by Thompson et al. [53] in their risk-score study and comprise Crohn’s disease (CD), multiple sclerosis (MS), psoriasis (PSO), rheumatoid arthritis (RA), systemic lupus erythematosus (SLE), and ulcerative colitis (UC). Type 1 diabetes (T1D) and celiac disease (CED) were excluded due to unavailable early reported data. Phenotypic information is also used to assess associations among different ADs through odds ratio (OR) analysis, with p-values determined via Fisher’s exact test in R, which also yields the OR and corresponding confidence interval. The phenotype in UKB can be retrieved using either the UKB ID or the ICD-10 code. The OR quantifies the strength of association between an exposure and an outcome, reflecting the likelihood of the outcome occurring in the exposed group relative to the unexposed group. An OR greater than 1 signifies comorbidity and a positive association, indicating that exposure increases the likelihood of the outcome. Conversely, an OR less than 1 suggests a lower comorbidity rate, implying a possible protective effect. An OR equal to 1 denotes no significant association between exposure and outcome.

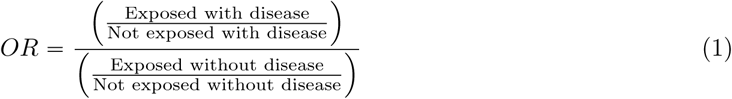

We utilize the earliest reported AD information from the UKB to categorize individuals by diagnosis in both the enhanced and standard datasets. Sample overlap is evaluated to determine the number of individuals shared across diseases and datasets, thereby defining the extent of cohort intersection and forming the basis for subsequent stratification analyses. Polygenic scores (PGS) from both datasets are then correlated to investigate pleiotropy, identifying groups of diseases with shared genetic architectures. The correlation analysis of risk scores originally conducted by Thompson et al. (2024) [53], with PGS values standardized according to their methodology. Given the standardized format of the PGS data, Pearson’s correlation coefficient is employed to quantify these associations.

A range of comparative analyses are performed, including two-dimensional polygenic score assessments that examine relationships between paired AD and display group divisions according to PGS levels, these approaches enhance interpretability. T1D and CED are omitted due to the absence of initial reported data from the UKB. To minimize noise from excessive data points and preserve clarity in pattern recognition, the enhanced dataset with fewer samples is used in this analysis. The association between PGS and disease categorization is visualized via scatter plots, generated using the ggplot2 library in R (version 4.4.1). All individuals from the refined dataset participate in the analysis except those with two ADs whose overlapping sample size is below 30; such cases are excluded to maintain the integrity of statistical analysis. Details pertaining to PGS were accessed on February 6, 2024.

### 2.2 Process for Detecting Genetic Variants in the Unrelated White British Subset of the UK Biobank and Linking Identified Genes to Biological Pathways

#### 2.2.1 Approach for reveling important Autoimmune Disease Genetic Variants and Conducting Their Functional Annotation

Following the analysis of the PGS datasets, the study proceeded to process the genetic data. For this purpose, variant related information was downloaded from Zenodo on May 30, 2024. Zenodo is an open-access digital repository that allows researchers to store, share, and preserve scientific outputs across various disciplines. The variant dataset, made available by Thompson et al. (2024) through Zenodo, was derived from a cohort of unrelated White British participants in the UKB. To determine significant variants, a genome-wide association threshold of *P* < −log_10_(5 × 10^−8^) (equivalent to a (minus log10 p ě 7.30103) was applied. The Human Leukocyte Antigen (HLA) region, located on chromosome 6, is notable for being both gene-rich and highly complex. Identifying the specific variants responsible for traits and diseases within the HLA region is often difficult because of its complexity. Consequently, this region was excluded from the analysis. Previous studies have shown that variants located near the HLA region tend to exhibit strong correlations due to their physical proximity, which can complicate the interpretation of genetic associations and obscure relationships with ADs [54]. Excluding this region and its flanking areas enhances the clarity of genetic analyses.

The dataset used in Thompson et al. (2024) was based on the GRCh37 assembly, and therefore the genomic coordinates for the HLA region correspond to the hg37 reference genome (28,477,797–33,448,354). To account for neighboring regions, a buffer of approximately 100,000 base pairs was added upstream and downstream, resulting in an excluded region from 27,400,000 to 34,400,000. Filtering out the HLA region reduced the number of variants available for most ADs. To improve genomic mapping accuracy and the relevance of disease associated variant studies, the coordinates were converted from GRCh37 to the latest human genome assembly, GRCh38, using the UCSC Lift Genome Annotations tool. This tool translates coordinates between assemblies, identifies genes with changed annotations, and utilizes pre-generated chain files for accurate conversion [55, 56]. The analysis was conducted between October 22, 2024, and February 25, 2025, using the “Dec. 2013 (GRCh38/hg38)” assembly.

The updated GRCh38 coordinates for single nucleotide polymorphisms (SNPs) were then used as input for the Variant Effect Predictor (VEP) via the Ensembl Genome Browser. VEP is widely employed to analyze, annotate, and prioritize variants across both coding and non-coding regions. For this study, GRCh38.p14 served as the reference genome, with VEP version v113.0 and the homo sapiens core 113 38 database. The analysis also incorporated data from the Genome Aggregation Database (gnomAD) version 4. The overall workflow is summarized in Figure 1.

**Figure 1:**
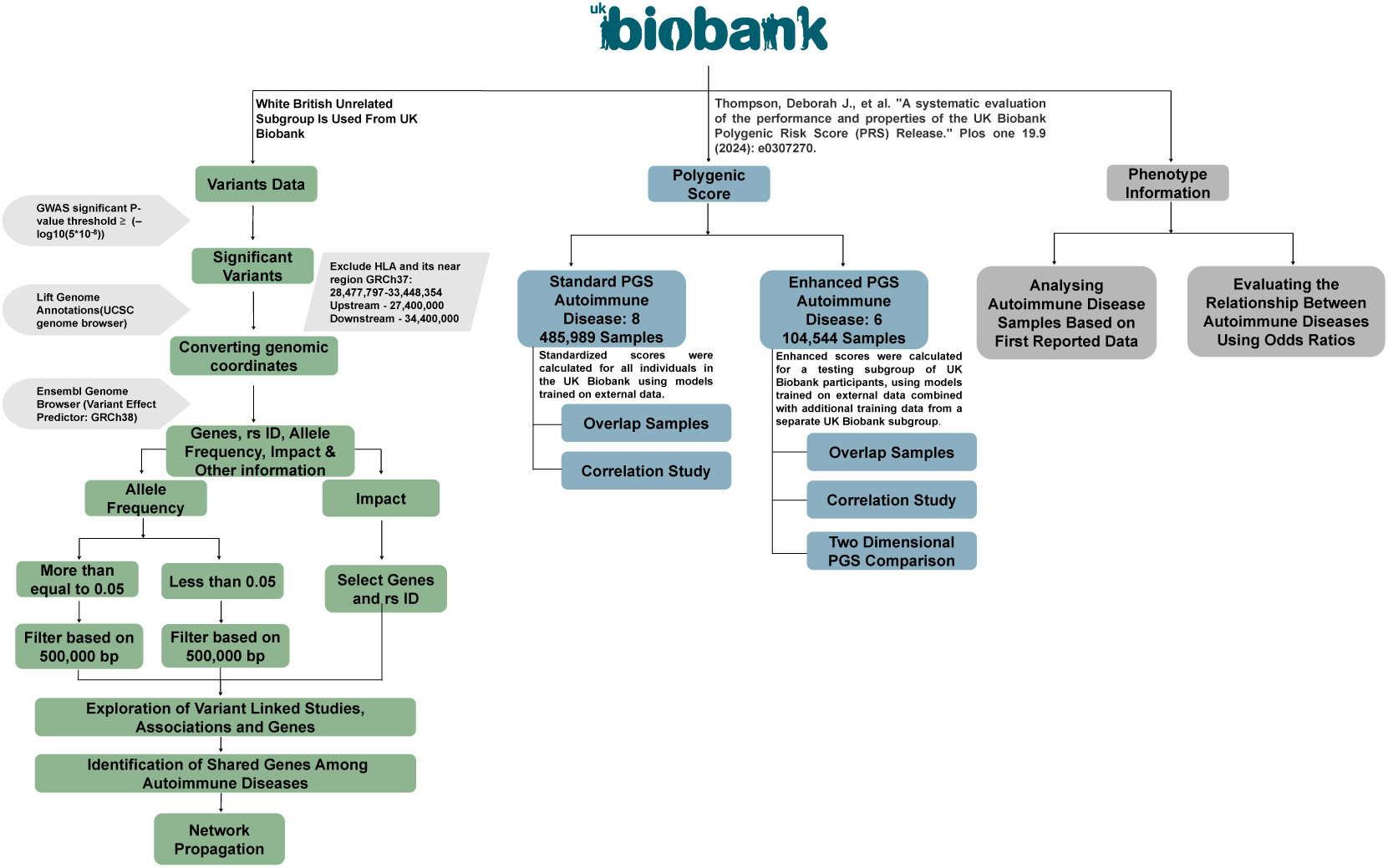
The flowchart outlines the overall study framework for analyzing data obtained from the UK Biobank (UKB). A color-coded scheme is used to distinguish the three categories of UKB data and their corresponding analytical procedures. The left panel (green) depicts the variant-level analyses conducted in the unrelated White British subgroup, emphasizing the detection of genes common to multiple autoimmune diseases (ADs). The central panel (blue) illustrates analyses of polygenic score (PGS) data, including correlation evaluations and examinations of risk score distributions. The right panel (dark gray) represents analyses involving phenotypic data, which are utilized to examine the co-occurrence of ADs and to characterize the study population. Sections in light gray indicate the software tools employed and the data extraction steps executed during the analysis. Further details regarding the data and analysis are provided in the “Method” section.

The VEP output provides information including chromosome location, variant position, reference and alternative alleles, beta value, standard error, -log10 p-value, and counts of cases and controls. The alternative allele represents the variant differing from the reference at a specific position, whereas the reference allele is not necessarily the most frequent in the population. Ensuring consistent reference/alternative allele coding across datasets is critical to correctly interpret the effect direction. Additionally, the −log 10p*p*q transformation of p-values is applied to facilitate their interpretation.

#### 2.2.2 Approach for Assessing Significant Variants Considering Allele Frequency and Functional Impact

To discover novel gene and variant connections pertinent to ADs, genetic variants are prioritized based on their allele frequencies and predicted functional consequences using annotations generated by the VEP tool. The analysis of VEP results involves two main filtering steps based on allele frequency (AF) and predicted functional impact. The AF metric, which represents the frequency of a particular allele in the population relative to all alleles for that gene, serves to differentiate common variants (AF ě 0.05) from rare variants (AF < 0.05). To account for the clustering of GWAS variants within linkage disequilibrium regions, variants situated within 500,000 base pairs of each other are grouped into loci separately for each AF category, generating two gene sets per AD. The functional impact of variants is assessed using VEP-assigned scores categorized as “high”, “moderate”, “low” or “modifier.” Most variants selected by AF filtering tend to have “modifier” or “low” impacts, often located in noncoding regions or causing minor amino acid substitutions unlikely to greatly affect protein function. A complementary filtering approach prioritizes variants based specifically on their predicted biological effects. Variants classified with high or moderate impacts, such as missense mutations, splice site variants, frameshifts, and in-frame deletions, are selected due to their greater potential to cause loss of function or significant changes in protein structure. This step produces a distinct set of genes linked to each AD. This methodology ensures robust identification of variants by integrating AF and functional consequence metrics to better understand genetic contributions to disease.

The final stage involves examining the rs-IDs and their associated genes using the GWAS Catalog to explore known genetic associations, relevant traits, and published studies. This comprehensive analysis integrates several layers of annotation and filtering based on unique rs-IDs, gathering both gene and transcript level data. Particular focus is placed on genes that are commonly identified across ADs through any of the applied methods, including rare variant analysis, common variant analysis, and predicted functional impact assessment. This approach enriches the interpretation by highlighting shared genetic factors.

#### 2.2.3 Network Propagation Strategy for Pathway Analysis of Genes Commonly Associated with Autoimmune Diseases

Network propagation analysis is conducted on genes shared by multiple ADs to enhance the genetic signal using existing interaction data. This method broadens the initial gene overlap into an expanded disease module by incorporating neighboring genes that likely participate in the same biological pathways, thereby revealing shared pathways and key hub proteins. For pathway exploration, this study utilizes hallmark gene sets from the Molecular Signatures Database (MSigDB), which represent well-defined biological processes and states characterized by coordinated gene expression patterns. These gene sets are computationally refined by identifying overlapping groups and retaining genes with synchronized expression, minimizing redundancy and noise. The curated collection, known as the H Collection in MSigDB [57], provides a biologically meaningful framework for pathway analysis widely used in research.

Network propagation starts with a predefined set of seed genes and utilizes the STRING v12 interaction network [58]. To ensure high confidence in interactions, only edges with score ≥ 700 are retained. Highly interconnected nodes, specifically those with degrees ≥ 900, and self-loops are removed, with the analysis confined to the network’s largest connected component. The diffusion of signals through the network is performed utilizing a regularized Laplacian kernel, which effectively smooths local neighborhood effects while dampening outliers. The initial raw scores obtained from this diffusion are then converted into permutation-based z-scores through 1,000 shuffles of seed genes, followed by standardization, which produces a reliable map of network-level effects.

All steps, executed within the R environment (version 4.4.1), utilize specialized libraries like diffuStats and GSVA (Gene Set Variation Analysis). This network-based approach aims to identify biological pathways functionally linked to genes with disease-associated variants. After filtering high confidence edges and removing hub nodes, the diffused signals are validated through multiple rounds of random seed shuffles, ensuring that the observed nodes are truly significant and not artifacts of noise. The final output presents a table of pathway scores ranging from –1 to +1; scores near +1 indicate strong association and activation, while those near –1 suggest pathway suppression. Scores around zero imply minimal network propagation.

### 2.3 Comparative Analysis of Autoimmune Comorbidity Patterns Using samples from UK Biobank and TriNetX

The AD selected for this analysis includes disease from different organ systems, such as neurological (MS, myasthenia gravis, inflammatory polyneuropathy, other demyelinating disease) to dermatological (vitiligo), gastrointestinal (CD, UC), multiple organ (other rheumatoid arthritis, ankylosing spondylitis [59], psoriatic and enteropathic arthropathies, SLE, reactive arthropathies) and internal organs (juvenile arthritis) and cutaneous (pemphigus, PSO)[60]. Fifteen ADs were included in this research, with analyses restricted to individuals of European ancestry/ white race. Each autoimmune disorder is characterized by a specific typical age of clinical onset; nonetheless, epidemiological studies have established that autoimmune conditions may manifest across the entire lifespan, without strict age boundaries [61]. The concept of age at onset denotes the time at which initial disease symptoms emerge for a given individual.

Within the TriNetX (TNX) database, the ’Age at Index’ variable represents the patient’s age at the occurrence of the primary studied event. Likewise, UKB provides disease-specific age indices for individual ADs. Accordingly, the age at onset was determined separately for each disease, accounting its unique clinical presentation and natural history. The data used from TNX is phenotypic data for different ADs. The data retrieval and analysis was done on May 30, 2024. Data from a global collaborative network with 120 healthcare organizations (HCOs) underwent analysis on the TNX platform. Defining cohorts through query criteria was the initial step. For each AD, the case cohort was defined as individuals with a confirmed diagnosis of the respective autoimmune condition and self-identified as white. Importantly, individuals with only control-related codes (ICD10CM:Z00) were excluded from the disease case group. Conversely, the control cohort consisted exclusively of white individuals with clinical encounters coded as ICD10CM:Z00, without any documented diagnoses of autoimmune disorders. Thus, no overlap of control or disease samples was permitted between case and control groups, ensuring the mutual exclusivity of the cohorts by both diagnostic and demographic criteria. The subsequent phase of analysis involves comparison of outcomes between cohorts. Propensity score matching was employed to select subjects in both case and control groups based on key covariates including sex, age, age at index event, and ethnicity. The TNX platform was utilized to compute OR for the development of comorbid conditions with AD across white race.

Phenotypic data from the UKB were obtained, with all relevant information downloaded on July 13, 2023, from the UKB official portal. The analysis was conducted under project ID 91246. A total of 502,371 samples were retrieved to calculate OR, spanning data for 15 ADs collected between 1902 and 2022. Multiple age cutoffs were applied to define the age at index for each disease. OR were computed using R studio with Fisher’s exact test. Comparative analyses of ORs derived from the two datasets were visualized using the R package “ggplot” for graphical representation and interpretation.

## 3 Result

### 3.1 Autoimmune Disease Sample Overlap and Comorbid Relationship Phenotype Based Evaluation

Frequent co-occurrence of autoimmune disorders highlights shared etiological mechanisms and steers mechanistic research directions. This section quantifies both the degree of sample overlap among different autoimmune conditions in the UK Biobank (UKB) and their comorbidity patterns. Overlap intensity is evaluated by tracking how often diagnoses are shared across patient cohorts, using first-diagnosed phenotype records from the UKB dataset. Phenotypic data is available for all 502,371 UKB participants. Among autoimmune diseases (ADs), psoriasis (PSO) has the largest sample count (15,978), followed by rheumatoid arthritis (RA, 13,344), and ulcerative colitis (UC, 6,702), whereas systemic lupus erythematosus (SLE) is represented by only 1,101 cases. Table 1 further displays that multiple sclerosis (MS) includes 2,593 patients, and Crohn’s disease (CD) comprises 3,503 cases. Suppl. Figure S1 visualizes both shared and unique patient counts, based on phenotype data. Each column of the figure marks a distinct set intersection, with dot patterns and set sizes reflecting disease associations. MS (2,367; 91.2%), PSO (14,379; 89.9%), and RA (11,528; 86.3%) have the highest numbers of independent (unshared) cases. Conversely, CD and UC exhibit the smallest proportions of unique cases, with (60.8%) 2,130 and (75.7%) 5,074 unshared samples, respectively. The percentage of unshared cases for each AD was calculated as the proportion of individuals with specific disease only relative to the total number of cases for that disease. The highest degree of sample overlap is observed between PSO and RA (993 shared samples), followed by UC and CD (922) and UC and RA (247).

**Table 1:** Summary of initial sample characteristics for selected autoimmune diseases (ADs) obtained from the UK Biobank (UKB). The table lists each disease with its corresponding ICD-10 code and UKB Field ID. It also presents the total number of identified cases and the distribution of male and female participants within the phenotype dataset.

| Diseases | ICD10 Code | Field ID (UKB) | No. of Patients (First Reported) | Male | Female |
| --- | --- | --- | --- | --- | --- |
| CD | K50 | 131626 | 3,503 | 1,568 | 1,935 |
| MS | G35 | 131042 | 2,593 | 721 | 1,872 |
| PSO | L40 | 131742 | 15,978 | 7,982 | 7,996 |
| RA | M05 | 131850 | 13,344 | 4,364 | 8,980 |
| SLE | M32 | 131894 | 1,101 | 166 | 935 |
| UC | K51 | 131628 | 6,702 | 3,358 | 3,344 |

Multiple combinations of ADs demonstrate positive ORs, several of which are statistically significant (p < 0.05). The computational approach for estimating the ORs is described in Section 2.1.1. The strongest positive associations are observed between SLE and RA (OR = 1.03), SLE and MS (OR = 0.58), and RA and PSO (OR = 0.45). A non-significant positive association is also identified between MS and PSO (OR = 0.06). In contrast, the association between MS and RA yields a slightly negative OR (–0.01), which does not reach statistical significance. The OR values were log10 transformed and presented in Figure 2, while the corresponding numerical data are provided in Suppl. Table S1.

**Figure 2:**
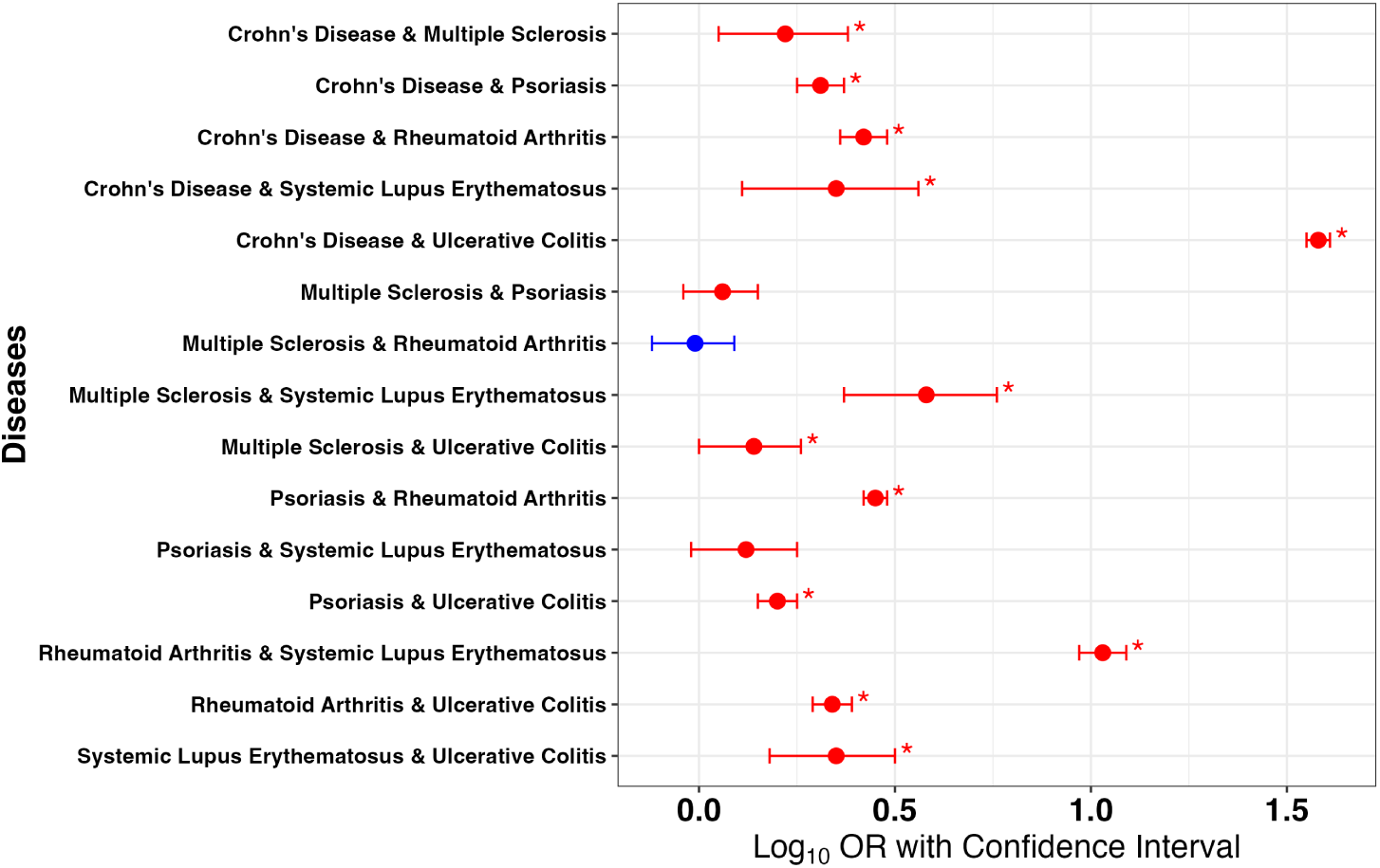
The forest plot presents the odds ratios for the comorbidity between autoimmune diseases. Each line corresponds to a specific pair of diseases, with the central area showing the log10-transformed odds ratio along with its confidence interval. Statistically significant associations (p < 0.05) are marked with a red star on the plot. The location of the odds ratio relative to the zero reference line indicates the nature of the association: values below zero suggest a protective effect, while values above zero indicate a positive comorbid relationship between the diseases.

### 3.2 Insights into Cohort Overlap, Risk Score Correlation and Distribution Across Autoimmune Diseases

Ensuring the validity of comparative studies and to avoid sample biases, cohort overlap within and between the standard and enhanced PGS datasets is investigated. The analysis incorporates participant data on ADs from both PGS datasets. Evaluating the extent of sample overlap and computing correlation measures across different phenotypes provides insights into shared genetic architectures and potential pleiotropic relationships, thereby enhancing the understanding of genetic risk patterns. The sample overlap comparison between the standard and enhanced datasets indicates that the enhanced dataset constitutes a subset of the standard dataset (Suppl. Figure S2). The two datasets share a total of 104,544 individuals, while the standard dataset has 381,445 unique samples. The enhanced dataset includes 6 ADs, while the standard dataset has 8 ADs, (Figure 1). Psoriasis (PSO) was the most prevalent condition in both the enhanced and standard datasets, with 3,142 and 15,527 patients respectively. Rheumatoid arthritis (RA) followed, with 2,747 patients in the enhanced dataset and 12,792 in the standard dataset. SLE had the smallest number of cases, 312 in the enhanced set and 1,054 in the standard, followed by MS with 478 and 2,488 cases respectively. Notably, SLE exhibits a significant sex imbalance, with 37 males and 275 females in the enhanced cohort, and 156 males alongside 898 females in the standard cohort (Table 2).

**Table 2:** The table presents patient numbers in both the enhanced and standard datasets for various autoimmune diseases. The first three columns specify the disease names, their ICD-10 classification codes, and the corresponding UK Biobank field identifiers. Subsequent columns detail the number of patients diagnosed with each condition. PGS data is available for all listed patients.

| Diseases | ICD10 Code | UKB ID | Enhanced Dataset |  |  | Standard Dataset |  |  |
| --- | --- | --- | --- | --- | --- | --- | --- | --- |
|  |  |  | Total | Male | Female | Total | Male | Female |
| CD | K50 | 131626 | 720 | 324 | 396 | 3,376 | 1,529 | 1,847 |
| MS | G35 | 131042 | 478 | 123 | 355 | 2,488 | 690 | 1,798 |
| PSO | L40 | 131742 | 3,142 | 1,580 | 1,562 | 15,527 | 7,805 | 7,722 |
| RA | M05 | 131850 | 2,747 | 842 | 1,905 | 12,792 | 4,206 | 8,586 |
| SLE | M32 | 131894 | 312 | 37 | 275 | 1,054 | 156 | 898 |
| UC | K51 | 131628 | 1,419 | 722 | 697 | 6,497 | 3,277 | 3,220 |

The pattern of sample overlap across diseases in both the enhanced and standard datasets is consistent with the phenotypic data. MS exhibits the largest proportion of unique or unshared samples, with 93.5% in the enhanced dataset and 91.3% in the standard dataset. RA follows closely, showing 85.9% unshared samples in the enhanced dataset and 86.4% in the standard. CD has the smallest proportion of unique cases, with 59.3% unshared in the enhanced dataset and 60.9% in the standard one. Ulcerative colitis (UC) ranks next, with unshared sample percentages of 76% and 75.9% in the enhanced and standard datasets, respectively (Suppl. Figure S3 and Suppl. Figure S4). Among diseases, PSO and RA share the highest number of overlapping samples, totaling 200 in the enhanced dataset and 952 in the standard. This is followed by CD and UC sharing 199 samples in the enhanced group and 884 in the standard.

All traits, diseases, and ADs are correlated using Pearson’s method. There are just a few unfavourable associations between diseases, traits, and ADs. Non-significant correlations, where the p-value exceeds 0.05, are indicated by a cross (x) symbol in Suppl. Figure S5 (enhanced dataset) and Suppl. Figure S6 (standard dataset). Within the standard dataset, several autoimmune disease pairs exhibit moderate positive correlations (above 0.4), such as RA and T1D with 0.40, SLE and CED with 0.42, and UC and CD with 0.54. Statistically significant negative correlations (below -0.2) are seen between MS and RA at -0.24, and T1D and MS at -0.22. The enhanced dataset reveals notable positive correlations (> 0.20) between RA and T1D (0.43), SLE and CED (0.39), and SLE and MS (0.21). Slight negative correlations occur between RA and MS (-0.28) and between RA and CED (-0.19). The strongest correlation above 0.50 is observed between UC and CD (0.53) in the standard set. Associations exceeding 0.40 include RA and T1D (0.43 in the enhanced, 0.40 in the standard set) and CED and SLE (0.41 in the standard set) (see Suppl. Tables S2 and S3).

### 3.3 Assessment of the Distribution of Genetic Risk Scores in the Enhanced Dataset

We compared the mean polygenic scores (PGS) for MS against those for PSO and RA (see Figure 3A and Figure 3B). Each point on the plot represents an individual patient’s risk score. The average PGS for MS is significantly different from that of both PSO and RA. The MS group’s mean score trends positively along the y-axis, while the averages for PSO and RA show positive trends along the x-axis. Most individuals fall within the central range of scores, indicated by a sharp peak for MS on the x-axis and for PSO and RA on the y-axis. The density plot along the x-axis for MS shows a slight peak shifted leftward, and PSO and RA demonstrate a slight rightward peak on the y-axis. Additionally, PSO and RA exhibit a wider distribution extending right on the x-axis, whereas the MS distribution along the y-axis spreads broadly toward the left. In both comparisons, patients with higher PGS cluster in the upper-right quadrant, producing a flattened distribution across both axes.

**Figure 3:**
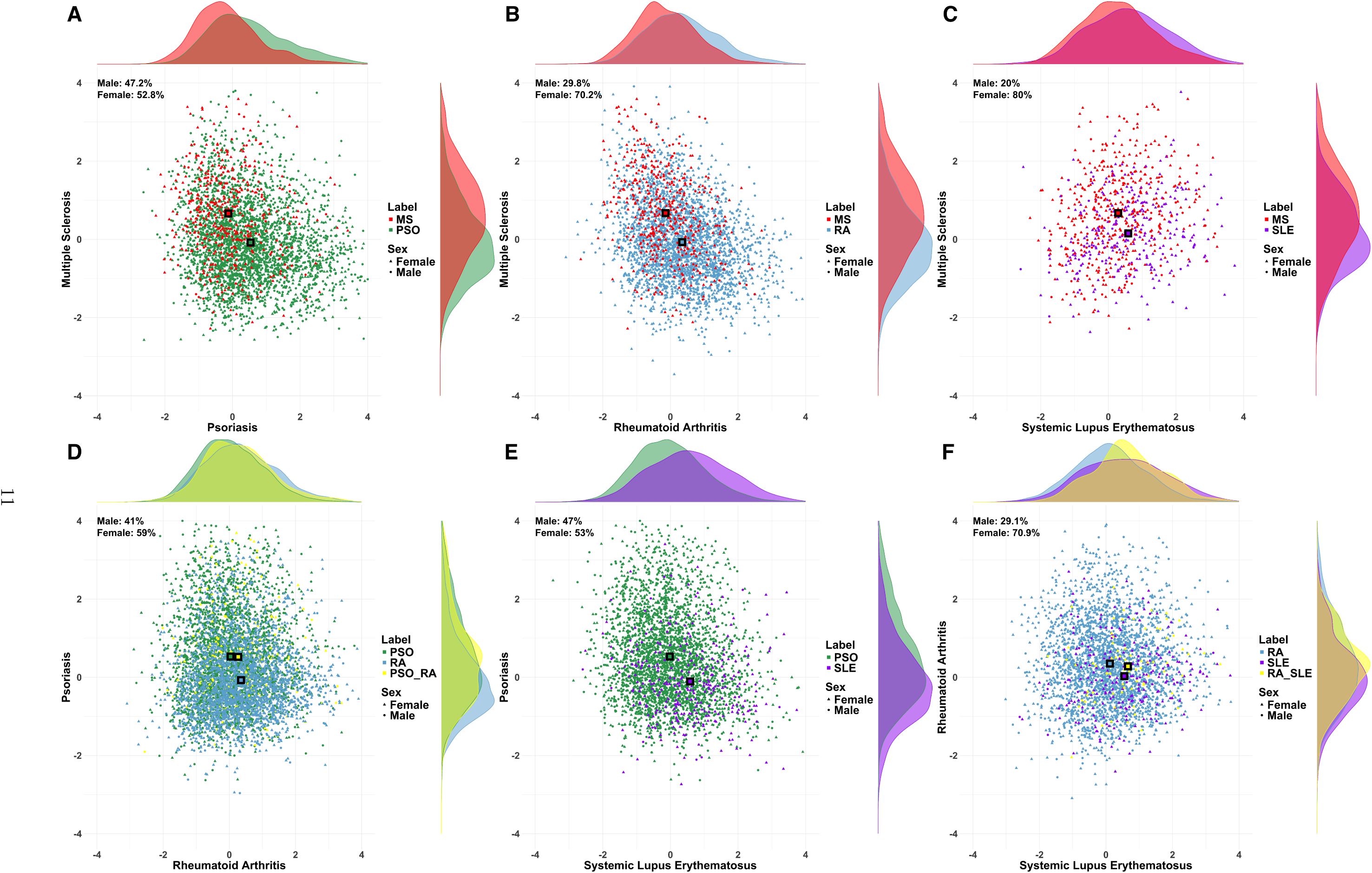
The scatter plot illustrates the relationship between the risk scores of two autoimmune diseases, revealing patterns and trends. Each point corresponds to an individual’s polygenic score (PGS) from the enhanced dataset, with distinct colors representing diseases: psoriasis (green; PSO), multiple sclerosis (red; MS), rheumatoid arthritis (blue; RA), and systemic lupus erythematosus (purple; SLE). Patients diagnosed with both diseases are highlighted in yellow. The axes span from -4 to 4. Data point shapes denote sex: circles indicate males and triangles indicate females. The proportions of male and female samples are indicated above each plot. Density plots alongside each axis display the distribution of data points across the range. Panels display the following comparisons: (A) PSO on the x-axis vs MS on the y-axis, (B) RA on the x-axis vs MS on the y-axis, (C) SLE on the x-axis vs MS on the y-axis, (D) RA on the x-axis vs PSO on the y-axis with dual-diagnosis samples labeled as PSO_RA, (E) SLE on the x-axis vs PSO on the y-axis, and (F) SLE on the x-axis vs RA on the y-axis with dual-diagnosis samples labeled as RA_SLE.

Furthermore, the distribution patterns of SLE and MS were examined. SLE exhibits a relatively uniform distribution with a slight rightward skew, whereas MS displays a higher density concentration toward the left side of the axis. Compared with SLE, fewer individuals with MS show lower PGS values along the x-axis. The peak density for SLE occurs in the lower range of the Y-axis, while MS demonstrates a broader distribution extending toward higher Y-values. Notably, both diseases share a considerable degree of overlap, indicating similar risk score values for most individuals. The observed pattern suggests a positive linear correlation between MS and SLE, with their mean PGS values trending upward along both the x and y axes (Figure 3C).

The subsequent pair of PSO and RA diseases exhibits divergent PGS profiles when examined individually, in contrast to the PGS indistinguishable diseases mentioned previously. Nevertheless, the risk associated with each disease is increased when these diseases coexist (Figure 3D). The density graphs on the x-axis display overlapping peaks for individual diseases, whereas the y-axis plot reveals a broader distribution for the comorbid group. The distributions of both individual disease groups are predominantly aligned with the comorbidity samples, with PGS values lying within the range observed for each condition. The RA peak on the y-axis indicates that certain individuals have relatively lower PGS values for PSO. The means of the two categories differ, despite the fact that the samples of PSO and RA are primarily overlapping. Conversely, samples with PSO demonstrate a positive mean for PSO but a negative mean for RA, whereas the mean of RA is positive for RA but negative for PSO. The mean of samples with both diseases is higher than that of samples with a single disease.

The comparison of PSO and SLE samples yields orthogonal mean locations, with each showing enhanced scores on one axis while remaining at baseline on the other. The PSO indicates a peak around the bottom of the x-axis; most people with this condition have lower PGS levels for SLE. Its distribution is more concentrated, with a smaller range of PGS values among these samples (Figure 3E). In comparison, the SLE distribution is wider and tilted toward higher PGS values, with a peak slightly to the right. The PSO distribution on the y-axis is larger, with a flatter curve at the upper end and more individuals with higher PGS values. The SLE peak indicates that some individuals have relatively low PGS values for PSO.

The blue and purple curves indicate samples with RA and SLE, respectively, whereas the yellow curve reflects individuals with both disorders (comorbidity) (Figure 3F). In the horizontal density plot (x-axis), the blue curve has a moderate peak towards the lower end, indicating that the majority of samples with RA cluster at lower x-axis values. The SLE curve has a wider dispersion and more variability in PGS along this axis. In the vertical density plot, RA peaks lower on the y-axis; samples generally have lower y-axis values. On the other side, the SLE has a little tilt toward higher values. The RA SLE curve, which represents samples with both disorders, peaks at the middle on both axes; these individuals have intermediate PGS values. (Detailed quadrant and gender distributions for all combinations are provided in Supplementary File.)

### 3.4 Identification of Genetic Variants in the Unrelated White British UK Biobank Cohort and Pathway Mapping of Associated Genes

#### 3.4.1 Significant Genetic Variants in Autoimmune Diseases and Their Functional Annotations

The variant data from Zenodo includes a large number of variants per AD, ranging from about 13 to 13.7 million SNPs (Table 3). RA has the highest variant count (13,628,693), closely followed by PSO and UC. SLE has the fewest variants (13,181,991), with T1D and MS also on the lower end. CD and CED fall in the mid-range. After excluding the HLA region and applying a GWAS threshold of *P* < −log_10_(5 ^ 10^−8^), significant variants reduce in number, detailed in the methods (2.2.2). Significant variants’ effect sizes (*β*) and standard errors are in Suppl. Table S4. CED shows the highest number of significant variants (4,228), followed by PSO, T1D, and SLE. MS, RA, UC, and CD have fewer significant variants (Suppl. Table S5). VEP output identified novel variants across ADs, with each assigned to genes (Suppl. Table S5).

**Table 3:** The table displays the number of variants utilized in the UK Biobank (UKB) polygenic score (PGS) analyses. Across different autoimmune diseases (ADs), the total variants/SNPs count is consistent, ranging between approximately 13.18 million and 13.63 million. Using a stringent p-value cutoff of 0.1, each disease involves around 2.7 million variants. When a more lenient threshold of 0.5 is applied, the number jumps to about 9.2 million variants, representing roughly a three to four fold increase. The differences in variant counts between diseases at each threshold are minimal, indicating a consistent distribution of association strength across ADs.

| Autoimmune Diseases | Abbreviation | <0.1 | <0.3 | Total |
| --- | --- | --- | --- | --- |
| Celiac disease | CED | 2,754,875 | 9,199,456 | 13,626,245 |
| Crohn's disease | CD | 2,753,381 | 9,238,161 | 13,618,476 |
| Multiple sclerosis | MS | 2,760,611 | 9,294,278 | 13,600,935 |
| Psoriasis | PSO | 2,741,332 | 9,181,598 | 13,628,692 |
| Rheumatoid arthritis | RA | 2,749,319 | 9,191,652 | 13,628,693 |
| Systemic lupus erythematosus | SLE | 2,718,449 | 9,205,720 | 13,181,991 |
| Type 1 diabetes | T1D | 2,726,237 | 9,280,203 | 13,544,480 |
| Ulcerative colitis | UC | 2,749,703 | 9,188,061 | 13,628,549 |

Filtering VEP output by common variants and 500 kb size reveals the most genes for PSO (15), CED (12), and T1D (7). CD and RA have the fewest (3 each), with SLE and UC intermediate 4 and 6 genes. After filtering rare variants, CED has the most associated genes (11), followed by SLE and T1D (9 each). PSO and MS each have six genes, CD three, and UC and RA four each. CED leads in genes with high/moderate impact variants (23), followed by T1D, SLE, and PSO, with the least in RA, CD, and UC. Table 4 and Suppl. Table S6 summarize genes identified under various filtering criteria across ADs.

**Table 4:**
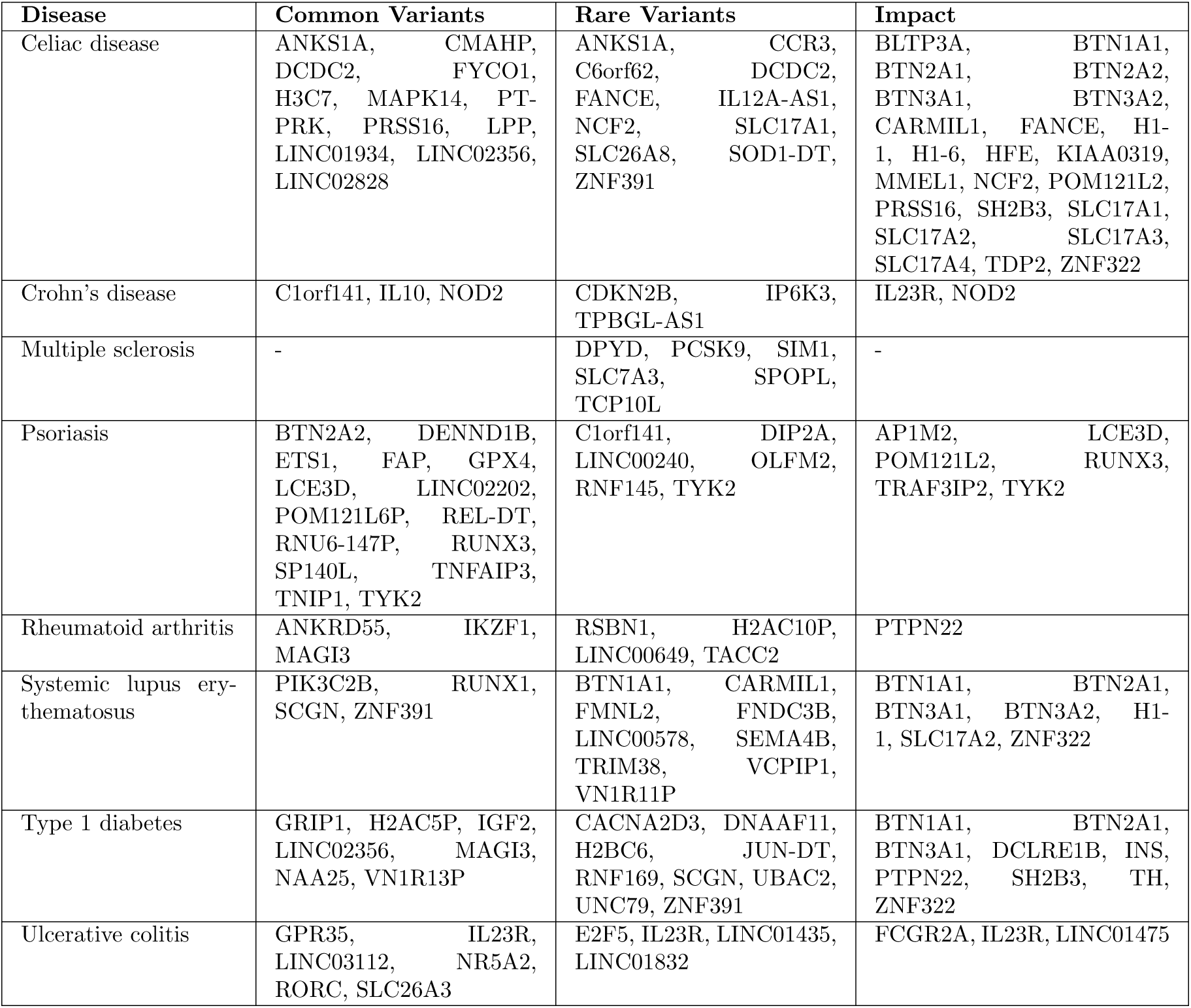
List of Genes Selected Based on Functional Impact and Variant Frequency.

#### 3.4.2 Identification of Shared Genes Across Autoimmune Diseases

This comprehensive comparison of gene sets related to variants selected by common, rare, and impact- based annotations from VEP output is inspired by the identification of genes shared across multiple ADs. By comparing these gene sets, we can identify those that consistently emerge across conditions, thereby offering insight into the core genetic drivers of autoimmunity. The ADs are associated with 14 genes. Out of these, 11 genes are associated with impactful variants, 2 have common variants, and 1 has a rare variant (Figure 4).

**Figure 4:**
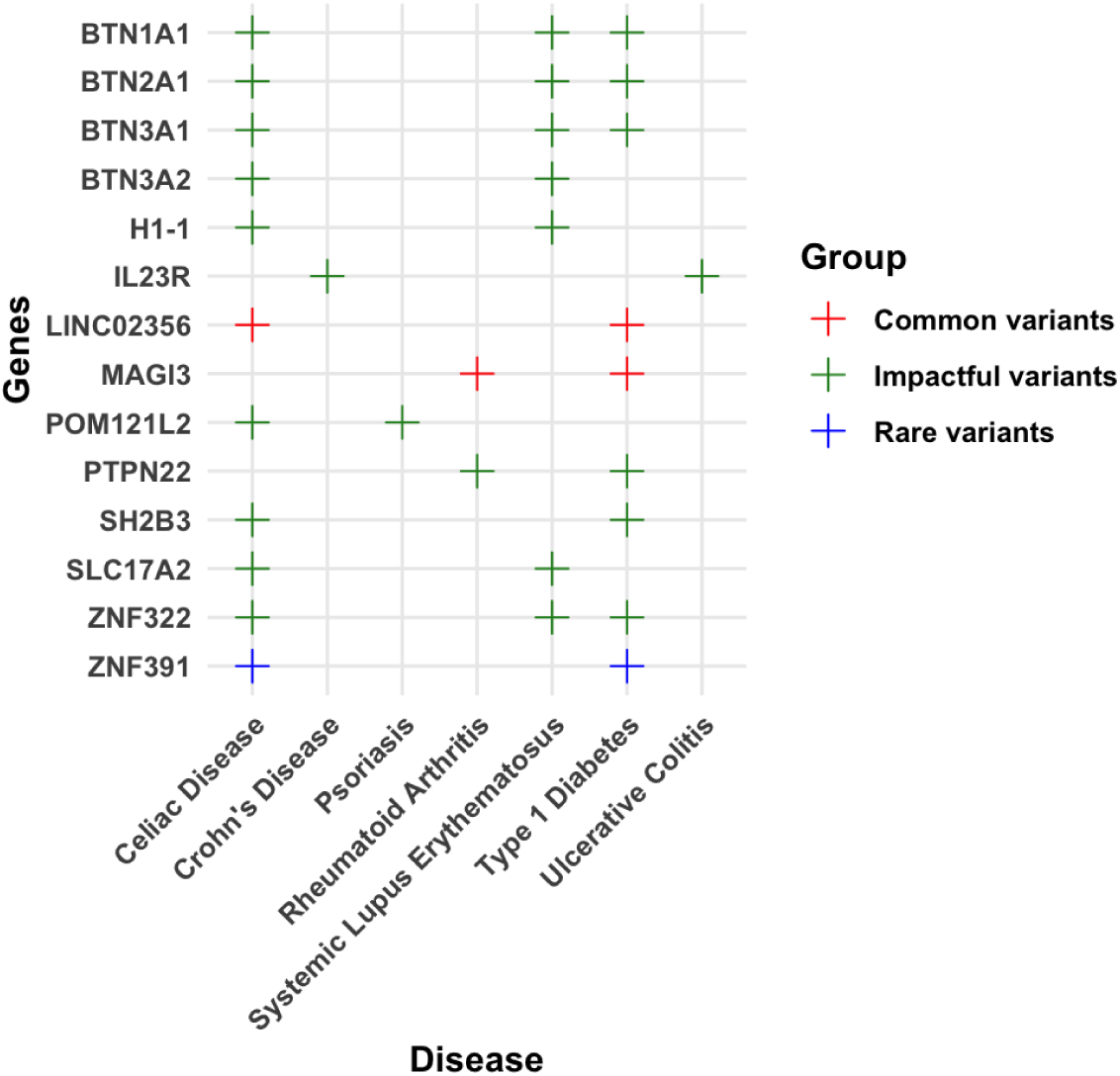
The plot shows genes shared among autoimmune diseases, with each gene color coded by variant type: blue for rare variants, red for common variants and green for high-impact variants. The x-axis lists the autoimmune diseases, and the y-axis indicates the common genes found across these diseases. Fourteen genes are identified as common to the autoimmune diseases, of which 11 are linked to impactful variants, 2 to common variants, and 1 to a rare variant.

Rare variants are commonly found in genes linked to T1D and CED. Variant rs67859638 in CED is associated with the gene ZNF391, showing a 2/2 association and trait in two studies, but it is not linked to any AD. Similarly, rs112328672 is connected to ZNF391 in T1D, yet no studies or AD associations are recorded for this variant. Both variants are located in an intron on chromosome 6 with a MODIFIER impact annotation. Although the ZNF391 gene has 19 associations and 13 traits reported in 17 GWAS catalog studies, none relate to ADs (Suppl. Table S7).

Three ADs (RA, T1D, and CED) share two genes containing three common variants. Variant rs10774624 appears in both CED and T1D, linked to 34 records across 26 traits in 33 studies, including associations with RA and vitiligo. Both variants are intronic/modifier and located in LINC02356 on chromosome 12, a gene with 75 associations and 53 traits reported in 64 studies related to RA and vitiligo. Variant rs1230666 is associated with RA, located in an intron of MAGI3 on chromosome 1, with 9/8 associations/traits in 9 studies but has no other autoimmune links or recorded studies. Similarly, rs72687973, an intronic modifier in MAGI3, is linked to T1D but lacks additional associations or studies. MAGI3 itself has 80 associations and 41 traits across 49 studies connected to CD, autoimmune thyroid disease, RA, T1D, and SLE.

The Impact-filtered dataset contains 21 unique variants across 11 genes on chromosomes 1, 6, 11, and 12 (Suppl. Table S7). BTN1A1 (chromosome 6) exhibits limited but significant overlap with CED, SLE, and T1D. The rs35555795 variant is a moderate-impact missense substitution found in all three diseases, supported by one study, with no other autoimmune disease links. BTN1A1 has 37 association records covering 25 traits across 34 studies for SLE and T1D and 40 associations with 27 traits in 37 studies for CED, without broader autoimmune connections. Additional moderate-impact missense variants rs3736781 and rs9393728 in BTN1A1 associate with CED in single studies and share similar gene-level metrics but lack further AD links.

BTN2A1 is commonly found on chromosome 6 in ADs CED, SLE, and T1D. Although gene-level data suggests possible links to RA and T1D, variant rs13195401 is consistently detected across all three diseases with eight associations over six traits documented in eight studies, but without additional AD connections. Variant rs13195402, also present in these diseases, shows higher associations for CED (nine associations and traits) and is linked to CD and IBD at variant and gene levels. It is associated with SLE and T1D in six studies with six associations but no specific AD references. Variants rs13195509 and rs3734542 exhibit similar patterns with fewer study associations (3 and 1, respectively), and no direct AD links despite gene-level connections to RA and T1D. Variant rs3734543 lacks variant-level association data but maintains gene-level autoimmune links with external database references. Overall, BTN2A1 shows a high number of trait associations (43/32 to 49/38 associations/traits) and studies (40 to 46), with recurring ties to RA and T1D at the gene level. BTN3A1 is a gene commonly associated with CED, SLE, and T1D. The rs41266839 variant, a moderate-impact missense mutation located on chromosome 6 for CED and SLE and chromosome 11 for T1D, appears consistently across these diseases. This variant has seven associations from seven studies, but none directly linked to ADs. BTN3A1 shows extensive associations at the gene level, with 30 associations across 22 traits in CED, 27/17 in SLE, and 28/17 in T1D, and is also connected to RA and T1D in several studies. The chromosome 6 variants rs13216828, rs71557335, and rs9358936 are frequently observed in CED and SLE and are linked to BTN3A2. Rs13216828 and rs9358936 are moderate-impact missense variants, while rs71557335 is a high-impact splice donor variant. While no variant-level association counts, study counts, or other AD linkages are specified for the individual variant. BTN3A2 shows multiple trait associations across studies in CED, 136–138 associations across 76 traits from 114 studies in SLE, 127 associations across 70 traits from 105 studies. Additionally, BTN3A2 is reported in the GWAS catalog as associated with RA. CED and PSO share the gene POM121L2, with two moderate-impact missense variants, rs16897515 and rs2235233, linked to CED. Rs16897515 associates with other ADs such as RA and T1D, supported by multiple studies, while rs2235233 lacks specific association data but shares gene-level links to RA and T1D. Variant rs41269255 in POM121L2 relates to PSO, supported by association studies but without known links to other ADs. The gene overall is linked to numerous autoimmune conditions and associated with multiple traits in studies. H1-1 on chromosome 6 is linked to both CED and SLE through the variant rs16891235, supported by three studies for each disease. Although this variant is associated with three studies and traits, it lacks broader AD connections at variant or gene levels. In contrast, IL23R on chromosome 1 relates to CD and UC via rs11209026, with 23 associations in 20 studies and a wide AD overlap. PTPN22, located on chromosome 1, shows the highest level of involvement across AD. IL23R is linked to multiple inflammatory and autoimmune conditions, including PSO, ankylosing spondylitis, RA, and IBD, with 71 studies covering 29 traits. Through rs2476601, it is associated with both RA and T1D, spanning over 100 variant-level associations in 102 studies and 48 traits. PTPN22 is also implicated in SLE, Graves’ disease, Hashimoto thyroiditis, and myasthenia gravis, with 109 studies reporting 54 traits in the GWAS catalog. SH2B3 (chromosome 12) is associated with CED and T1D via rs3184504, a variant with extensive study and association counts (>250), and also links to MS, autoimmune thyroid disease, and autoimmune hepatitis. At the gene level, CED shows a single association. For T1D, the GWAS catalog lists 395 associations across 201 traits from 362 studies. SLC17A2 and ZNF322 (chromosome 6) are observed in CED, SLE, and T1D, though without variant-level data. SLC17A2 shows 70 associations in CED (47 traits, 69 studies) and 63 in SLE (42 traits, 62 studies), with additional links to PSO. ZNF322 demonstrates involvement in CED (84 associations, 45 traits, 65 studies) and T1D (71 associations, 38 traits, 55 studies), but none for SLE.

#### 3.4.3 Network Propagation Identifies Key Biological Pathways Related to Common Identified Genes

The lists of seed genes used for network propagation analysis is presented in Figure 4. Pathway enrichment analysis reveals both common and unique biological mechanisms across ADs. The IL6- JAK-STAT3 signaling pathway is strongly upregulated in CED, CD, T1D, and UC but downregulated in PSO. Cell proliferation pathways, including the G2M checkpoint and E2F targets, are activated in CED but suppressed in T1D. PSO exhibits downregulation in immune, metabolic, and developmental pathways, whereas T1D shows activation in immune, cellular component, and developmental pathways. We focused on 15 AD related pathways, categorized into immune response, metabolism, developmental processes, signal transduction, cellular components, and fundamental biological pathways shown in Figure 5. Network propagation scores are provided in Suppl. Table S8, with additional explanations in the suppl. file, and all Hallmark pathways visualized in Suppl. Figure S7.

**Figure 5:**
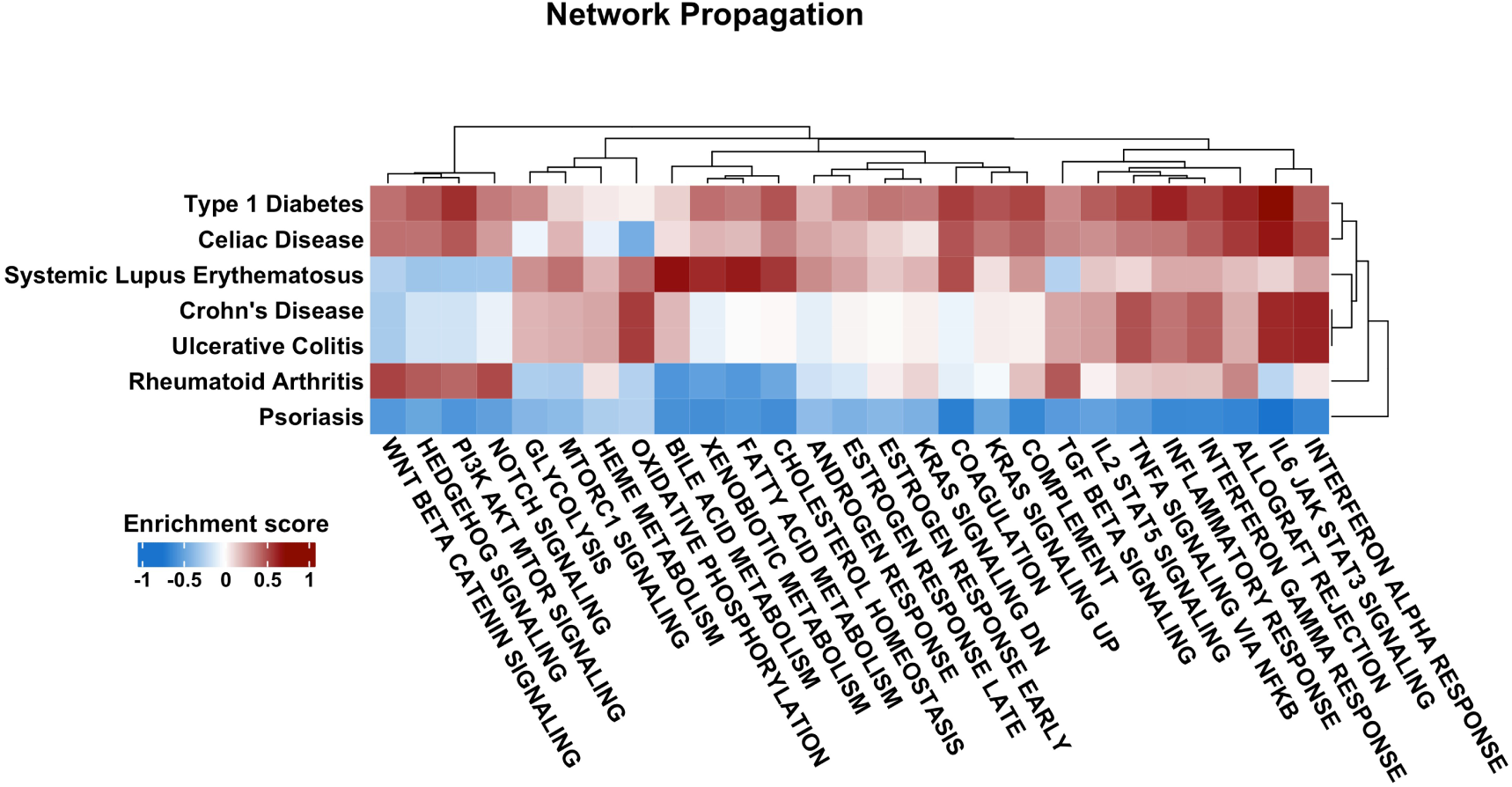
The heatmap shows a subset of the network propagation analysis, illustrating associations between autoimmune diseases and biological pathways identified through rare, common, and impact-based variant filtering. It focuses on 15 AD-related pathways, with rows representing pathways and columns representing diseases. Hierarchical clustering with dendrograms depicts interrelationships, and the color scale (–1 to 1) reflects enrichment magnitude, where positive values indicate stronger variant-driven contributions to disease mechanisms.

## 4 Comparison of Autoimmune Comorbidity Patterns Across UK Biobank and TriNetX

Comorbidity observation in TriNetX (TNX) and UKB relies on ORs derived from ADs. Most disease terms are consistent across both databases, except for two diseases that have different names. To standardize terminology, a mapping table linking disease names to common identifiers like ICD codes can be created. We compared ICD10 codes and disease names between the databases to ensure consistency. For example, TNX refers to myasthenia gravis and enteropathic arthropathies as ”myasthenia gravis and other myoneural disorders” and ”psoriatic and enteropathic arthropathies” respectively in UKB; however, their ICD10 codes match (G70.0 and M07; Suppl. Table S9). For further analyses, we use the UKB terms to maintain consistency. One discrepancy exists for postinfective/reactive arthropathies where TNX uses ICD10 code M02, while UKB uses M03.

Data from 75 to 80 of the 120 healthcare organizations in the TNX network included 163,606,965 patients, with 71,069,654 identified as white. Demographic characteristics across AD cohorts were analyzed, assessing age ranges, means, standard deviations, sex distribution, and cohort sizes (Suppl. Table S10). Baseline characteristics of patients before and after propensity score matching are summarized in Table 5. The average age of disease onset ranged widely from 1 to 90 years, averaging 65 years. Ankylosing spondylitis (53%), inflammatory polyneuropathy (54%), and postinfective/reactive arthropathies (51%) were more common in males, whereas juvenile arthritis (70%), MS (73%), demyelinating diseases (66%), RA (72%), and psoriatic and enteropathic arthropathies (63%) predominated in females.

**Table 5:** Participant Characteristics Pre and Post Propensity Score Matching.

| Characteristics |  | Before propensity matching | After propensity matching |
| --- | --- | --- | --- |
|  |  | White Race |  |
| Current Age | Patient Cohort | 6,697 | 6,697 |
|  | Control Cohort | 8,154,296 | 6,697 |
| Female | Patient Cohort | 3,781 | 3,781 |
|  | Control Cohort | 4,413,111 | 3,774 |
| Male | Patient Cohort | 2,913 | 2,913 |
|  | Control Cohort | 3,727,875 | 2,920 |

Comorbidity analysis of 15 ADs showed “other rheumatoid arthritis” with the highest patient count (395,932), followed by PSO (320,554), UC (177,016), and CD (173,481). Less common diseases included psoriatic and enteropathic arthropathies (4,527), pemphigus (5,593), postinfective/reactive arthropathies (9,137), and juvenile arthritis (29,413).

The UKB dataset included 502,371 participants (229,069 males, 273,302 females). PSO was most prevalent (13,110), followed by other rheumatoid arthritis (12,708) and UC (6,522). Smaller numbers were observed for postinfective/reactive arthropathies (14), pemphigus (51), juvenile arthritis (65), and myasthenia gravis (416). Ankylosing spondylitis showed the strongest male predominance (68%), while SLE was predominantly female (84%). Participant birth years ranged from 1934 to 1970, with age filters applied based on disease onset to refine cohorts (Suppl. Table S11).

The ORs from both databases show similar trends, with certain diseases exhibiting very high and statistically significant ORs (> 20), reflecting strong associations. MS and other demyelinating diseases have the highest ORs in the UKB (206.2) and TNX (78.5) datasets. Following these are ankylosing spondylitis with juvenile arthritis (UKB: 39.8, TNX: 23.7), other demyelinating diseases with inflammatory polyneuropathy (UKB: 39.1, TNX: 23.1), and juvenile arthritis with other rheumatoid arthritis (UKB: 21.1, TNX: 66.5). Some notable differences in ORs, although still significant, were found among dermatological and gastrointestinal diseases, such as PSO with MS (UKB: 1.18, TNX: 0.51), inflammatory polyneuropathy (UKB: 2.04, TNX: 0.74), myasthenia gravis (UKB: 1.59, TNX: 0.75), UC with MS (UKB: 1.43, TNX: 0.57), and other demyelinating diseases with myasthenia gravis (UKB: 2.45, TNX: 0.81), as detailed in Suppl. Table S12.

For multi-organ ADs such as psoriatic and enteropathic arthritis and other rheumatoid arthritis, ORs exceeded one in both the UKB and TNX datasets; however, significance was noted in only one database for certain associations. For example, psoriatic and enteropathic arthropathies had elevated but non-significant ORs with demyelinating diseases, inflammatory polyneuropathy, and vitiligo in TNX, whereas these associations were significant in UKB. Conversely, some comorbidities showed significant ORs in TNX but not in UKB, including RA with other demyelinating diseases and pemphigus, ankylosing spondylitis with other demyelinating diseases, postinfective/reactive arthropathies with CD, and psoriatic/enteropathic arthropathies with myasthenia gravis. Some ORs were zero in UKB, indicating no co-occurrence observed, with TNX similarly reporting missing ORs due to absence of relevant control cases. The broad range of ORs (0 to 206) was log-transformed for visualization (Suppl. Figure S8).

Differences between cohorts included reversed OR directions for several disease pairs. For instance, demyelinating diseases with PSO, MS with UC, myasthenia gravis with PSO, inflammatory polyneuropathy with PSO, CD with vitiligo, and MS with PSO showed negative log ORs in TNX (suggesting inverse or protective relationships) but positive in UKB (indicating increased risk), both statistically significant. MS and PSO had a strong inverse association in TNX (Log OR = -0.66) compared to a weak positive association in UKB (Log OR = 0.17). TNX tended to display stronger negative associations involving PSO, UC, vitiligo, and MS, while UKB showed milder or reversed trends toward positive associations. Consistent positive correlations were found for CD with UC (TNX: 3.06; UKB: 3.66) and MS with other demyelinating diseases (TNX: 4.36; UKB: 5.33), reflecting shared risk factors. Variations in effect sizes, such as between psoriatic and enteropathic arthropathies and PSO (TNX: 2.02 vs. UKB: 5.11), and non-significant associations like CD with pemphigus, likely reflect population, environmental, or biological differences.

## 5 Discussion

### 5.1 Informative Insights into Phenotypic Convergence and Comorbidity Dynamics in Autoimmune Disorders

Phenotypic sample overlap analysis revealed both shared and distinct patterns among autoimmune diseases (ADs). Substantial non-overlap highlighted phenotypic and etiological uniqueness, whereas overlap suggested convergent immune pathways. These patterns inform inter-disease relationships and molecular distinctions. Although Crohn’s disease (CD) and ulcerative colitis (UC) are distinct entities, rare co-occurrences such as one patient with both over 11 years and sisters affected within a year suggest partial immune overlap [62, 63]. Sample overlap likely reflects shared mechanisms and diagnostic uncertainty, as those with mixed symptoms may be labeled as having indeterminate colitis. Variation in UK Biobank (UKB) sample sizes among ADs likely arise from epidemiological and methodological factors. Common diseases like rheumatoid arthritis (RA) and psoriasis (PSO) are overrepresented due to higher prevalence, whereas systemic lupus erythematosus (SLE), CD, and multiple sclerosis (MS) are underrepresented owing to earlier onset relative to UKB’s recruitment age (40–69 years). Most AD pairs showed positive, significant OR associations, consistent with widespread comorbidity driven by shared genetic and immune factors. Positive but non-significant associations reflected limited power or divergent origins, and inverse associations were absent, underscoring strong interconnectivity among ADs and the importance of considering comorbidity in research and clinical settings. These findings mirror epidemiological evidence from Conrad et al. (2023) [4], who reported frequent comorbidity among connective tissue diseases (SLE) but minimal overlap for MS. This heterogeneity parallels our strong polygenic score (PGS) correlations for SLE–MS and RA–PSO pairs, versus weak or inverse correlations for others. Similarly, Chen et al. (2024) [64] showed that multi-phenotype PGS integration improves prediction accuracy, supporting multi-trait modeling to capture shared autoimmune mechanisms.

### 5.2 Interpretation of Correlations and Genetic Risk Patterns in Autoimmune Diseases Using PGS

The analysis of correlations between risk scores and disease presence reveals important shared biological mechanisms. Type 1 diabetes (T1D) and RA, as well as SLE and celiac disease (CED), show significant positive associations, indicating common immunological and genetic factors contributing to their co-occurrence. These links are supported both by clinical phenotype data and PGS correlations, highlighting overlapping genetic architecture. Multiple sclerosis (MS) and SLE exhibit a strong comorbidity with a high odds ratio (OR) and positive PGS correlation, suggesting a shared genetic susceptibility. Similarly, PSO and RA have a positive OR and modest PGS correlation, indicating partial genetic overlap. In contrast, PSO and SLE demonstrate a non-significant positive OR but negative PGS correlation, implying that while clinical features may overlap, their genetic backgrounds are distinct and potentially oppositional.

Distinct risk-score patterns for MS, PSO, SLE, and RA illustrate how genetic and environmental factors shape unique disease profiles. These risk scores reflect largely independent but partially shared genetic architectures, consistent with prior studies showing limited overlap among these autoimmune disorders. The data reveal a modest but meaningful inverse genetic relationship, notably between MS and PSO or RA, where individuals with higher genetic risk for MS tend to have lower risk for PSO [65] and RA [66], reinforcing a weak negative genetic correlation. This is also seen in the positive trend between MS and RA. Overall, the risk score distributions support the notion of distinct genetic risk profiles for MS and RA despite their shared classification as ADs.

Although ADs show opposing average PGS by clustering in different quadrants, over half of the samples spread across all quadrants for each disease pair, indicating substantial overlap in individual genetic risk profiles. This suggests that despite modest differences in average risk, the underlying genetic architectures are not completely separate. The wide distribution implies many individuals carry genetic susceptibility to multiple ADs, supporting the concept of polyautoimmunity and shared genetic predisposition. Thus, while quadrant averages reflect general trends, the broad sample overlap highlights the complex and partly shared genetic basis of autoimmune disease risk.

The near-orthogonal mean PGS of PSO and SLE indicate minimal genetic similarity [67]. SLE samples cluster mainly in the bottom-right quadrant (44%), showing a polarized pattern with higher scores on the X-axis, while PSO samples distribute more evenly, reflecting greater individual variability without clear directional bias. This supports their genetic divergence despite some overlapping clinical traits. In contrast, SLE and MS show a moderate shared genetic risk [68], with mean PGS values trending positively on both axes and a concentration of samples in the top-right quadrant, highlighting significant overlap in individuals with elevated risk for both diseases. Although over half of samples differ in risk distribution, the alignment of means and dense overlap suggest a meaningful genetic correlation and possible comorbidity between MS and SLE.

The PGS distribution for PSO and RA illustrates both shared and distinct genetic risks, shedding light on their comorbidity. Despite overlap, their group means diverge, with PSO samples showing high PSO but low RA risk and RA samples exhibiting the opposite, indicating partially independent genetic backgrounds. Individuals with both conditions have elevated PGS for each, reflecting an additive genetic burden. The quadrant analysis reveals complexity: while many PSO samples spread across quadrants with varying RA risk, RA samples cluster mainly in the bottom-right quadrant, indicating high RA and low PSO risk. Most individuals with both diseases fall in the top-right quadrant, consistent with high genetic risk for both. These results highlight the genetic heterogeneity in autoimmune comorbidity, emphasizing both shared and disease-specific loci.

The PGS analysis for SLE and RA shows that individuals with both diseases typically have moderate genetic risk for each [69], suggesting that comorbidity arises from overlapping but not extreme genetic factors. SLE exhibits greater variability in genetic risk compared to the more concentrated RA distribution. Most comorbid cases cluster in the high-risk quadrant for both diseases, indicating shared genetic influences. Overall, these results emphasize the complex, partially overlapping genetic basis of RA-SLE comorbidity.

More broadly, over half of the PGS distributions across autoimmune disease pairs lack clear, uniform patterns, highlighting the genetic complexity and heterogeneity of autoimmunity. Shared genetic factors, especially within the HLA region, can produce correlated or overlapping PGS distributions that mask distinctions between diseases. Additionally, comorbidities, diagnostic overlaps, and environmental and epigenetic factors contribute to further variability.

The clear female predominance in ADs, with about 60% of cases occurring in women, highlights the important role of sex-based biological factors in AD susceptibility. This is especially true for diseases like SLE and RA, where hormonal effects, primarily estrogen’s immune-activating properties, increase immune responsiveness in females [70, 71]. Furthermore, genes on the X chromosome and incomplete X-chromosome inactivation can lead to higher expression of immune-related genes, increasing women’s vulnerability to autoimmune conditions [72]. In contrast, PSO exhibits a more balanced sex ratio, suggesting a different disease mechanism less influenced by X-linked factors [73].

### 5.3 Integrative Insights into Genetic Architecture and Shared Gene Functions in Autoimmune Disorders

After exclusion of the HLA region, PSO, T1D, and CED retain a substantial number of risk variants. This persistence likely reflects their highly polygenic nature, strong non-HLA variant associations, and relatively homogeneous genetic profiles. Conversely, RA, SLE, UC, and CD display a stronger dependence on HLA-linked loci, resulting in a notable decline in detectable associations once this region is removed. These differences emphasize the necessity of accounting for disease-specific genetic architectures when interpreting results from Genome-wide association studies (GWAS) and PGS analyses. The marked decrease in significant variants for MS after HLA exclusion further indicates that its genetic susceptibility is highly concentrated within this locus, reinforcing antigen presentation as a key pathogenic pathway. Variant annotation helps enrich for biologically meaningful effects but inevitably omits variants situated in extended linkage disequilibrium regions or distant regulatory elements. Although this approach enhances interpretability and biological plausibility, it simultaneously reduces the total number of variants assessed for both common and rare categories. The observed enrichment of high- and moderate-impact genes in CED, T1D, SLE, and PSO suggests a greater functional variant load, whereas RA, CD, and UC show fewer such genes, possibly reflecting distinct underlying genetic mechanisms.

We explored the genetic landscape of ADs by identifying gene variants through comprehensive filtering of common, rare, and functionally impactful alleles. The notable scarcity of genes defined exclusively by common or rare variants highlights the importance of analyzing the full range of variant frequencies. Among 14 shared genes, 11 were identified based on functional impact, indicating that predicted effect size is a more reliable marker of shared autoimmune risk. A rare variant shared between type 1 diabetes (T1D) and CED, located in a gene not previously linked to autoimmunity in the GWAS Catalog, illustrates how infrequent but functionally significant alleles can reveal novel mechanisms underlying ADs. Its low frequency likely limits its detection in large-scale GWAS that focus primarily on common variants, explaining the underrepresentation of rare variant associations in autoimmune research.

This analytical framework enabled the discovery of both established and previously unreported gene disease associations. Several genes such as IL23R, PTPN22, SH2B3, and MAGI3 have already been documented in the GWAS catalog as being linked to multiple autoimmune disorders. Their re-identification in this study serves to validate the methodological approach and confirm the reliability of the analytical approach. In contrast, genes including ZNF322, POM121L2, and LINC02356 emerged as novel candidates to new ADs, likely reflecting the detection of variants not previously described. Although some of these genes have been implicated in individual ADs, the current results reveal new associations across additional conditions, thereby broadening their known biological significance. Furthermore, BTN1A1, H1-1, and ZNF391 have not been previously connected to autoimmune pathology in either published literature or GWAS databases; the present findings provide new evidence supporting their potential involvement.

These novel associations emphasize the importance of functionally diverse variants including intronic, non-coding transcript, missense, and splice-site variants, that may modulate gene regulation, splicing dynamics, or protein activity. Unlike conventional GWAS, which tend to emphasize exonic and common variants, the impact-based filtering strategy employed here facilitated the identification of biologically meaningful variants with broader functional implications, extending beyond the limits of standard association studies.

Additionally, the detection of multiple butyrophilin (BTN) family genes BTN2A1, BTN3A1, BTN3A2, and BTN1A1 across distinct ADs suggests a more extensive immunomodulatory role for this gene family than previously recognized [74]. The re-identification of MAGI3, which is known to participate in autoimmune pathways but lacks detailed variant-level annotation, underscores existing gaps in current GWAS datasets and illustrates how functional variant annotation can help bridge these gaps and refine our understanding of autoimmune pathogenesis.

#### 5.3.1 Network Based Pathway Mapping of Common Genes for Uncovering Biological Mechanisms

The network propagation outcomes illustrate how genetic risk variants shape disease biology through interlinked functional pathways. Applying strict filtering retaining only high-confidence connections, excluding central hubs, and using Laplacian diffusion with permutation testing greatly reduces random noise. Consequently, the proteins that remain statistically significant are likely to represent genuine functional associations rather than artefacts. Nonetheless, independent replication or functional assays are required to establish biological causality. Positive enrichment patterns observed in disorders such as CED, SLE, and UC indicate broad activation of interconnected pathways. This suggests that risk genes in these diseases occupy central network positions, influencing multiple immune, developmental, and metabolic processes [75, 76]. Such profiles are consistent with extensive, system-wide dysregulation. Conversely, PSO shows marked negative enrichment, implying a more localized or repressive network effect, reflecting pathway downregulation or silencing consistent with prior evidence [77]. The contrasting enrichment directions positive versus negative likely correspond to different pathogenic modes: hyperactivation in some diseases (e.g., inflammation or cellular proliferation) versus suppressed activity in others (e.g., reduced metabolism or immune tolerance). Shared positively enriched pathways such as IL6–JAK–STAT3 signaling point to common autoimmune mechanisms, while distinct propagation profiles reveal disease-specific biological features. The network propagation analysis provides insight not only into which pathways are affected but also into how disease-associated genes interact within broader molecular systems either amplifying or dampening network activity thus adding a functional dimension to genetic association results.

### 5.4 Integrative Interpretation of Comorbidity Patterns Across TriNetX and UK Biobank Cohorts

Differences in terminology and ICD-10 coding between TNX and UKB pose major challenges to inter-operability and data standardisation, potentially leading to cohort misclassification and biased analyses that affect epidemiological and clinical interpretations. Sex-based disparities in AD prevalence within the White UKB population higher ankylosing spondylitis in men and increased MS, RA, and other demyelinating diseases in women underscore the importance of sex as a biological variable in AD pathogenesis, consistent with prior studies [78]. Factors such as oestrogen-mediated immune modulation, X-linked genetics, and diagnostic bias may contribute to these trends. Strong associations between MS and other demyelinating diseases, and between ankylosing spondylitis and juvenile arthritis, corroborate previous evidence of shared genetic and immunological mechanisms [79, 80]. The frequent clustering of autoimmune and inflammatory conditions suggests overlapping biological pathways driven by genetic predisposition, immune dysregulation, and environmental influences. Discrepancies in ORs for certain dermatological and gastrointestinal diseases—exceeding 1 in UKB but below 1 in TNX likely reflect differences in cohort composition, diagnostic criteria, and data collection methods [81], as well as sample size and inclusion criteria. Variations in multi-organ autoimmune disease associations further warrant detailed investigation. The broader but more heterogeneous TNX cohort yielded non-significant ORs (>1) due to unmeasured confounders and data granularity, whereas the deeply phenotyped UKB produced significant associations through stricter covariate control but limited case numbers. Divergent sex ratios and ascertainment methods between datasets likely contributed to these differences. Some disease pairs showed opposing log-odds protective in TNX but risk-enhancing in UKB indicating that genetic architecture, environment, and cohort characteristics distinctly influence associations. Consistently elevated ORs across both datasets, however, support shared etiological pathways and robust biological links. These findings emphasise the need for harmonised analytical frameworks, standardised disease definitions, and larger integrated cohorts to enhance reproducibility and cross-dataset comparability in AD research.

## 6 Limitations

This study faces several limitations related to dataset structure, disease representation, and analysis methods in autoimmune disease (AD) research. Unequal representation of ADs in the UK Biobank (UKB) arises from case identification relying on hospital records, primary care data, and self-reports, each with differing accuracy. Complex symptoms and diagnostic delays can cause misclassification, leading to sample sizes that may not mirror true disease prevalence, which affects statistical power and generalizability. Comorbidity assessments based on odds ratios only capture concurrent diseases, not causality or temporal relationships. Polygenic score (PGS) correlations show genetic overlap but lack functional interpretation and are influenced by sample size imbalances, sex biases (e.g., female predominance in SLE), and overlapping cases with differing genetic profiles, complicating risk interpretation. PGS calculations mainly involve GWAS from European ancestry, limiting cross-population use, and reflect additive effects of common variants, excluding rare variants, gene interactions, and environmental factors involved in autoimmunity.

Variant-level analysis depends on computational predictions requiring experimental validation; rare variant analysis is statistically challenging due to low frequencies. Some newly associated genes lack known autoimmune relevance, making functional interpretation speculative. The GWAS Catalog only includes genome-wide significant variants, possibly missing related but less prioritized variants. Enrichment using MSigDB hallmark gene sets, though biologically informative, misses context-specific functions and assumes static gene-pathway links. Network propagation highlights pathways but does not identify causal genes and may inflate signals due to network structure. Restricted access to raw data limits reproducibility. Additionally, the UKB cohort is healthier, predominantly European, and skewed towards older middle-aged volunteers, limiting generalizability. Baseline and self-reported data are static, prone to recall bias, with missing phenotypes reducing analysis depth. Ethical and regulatory challenges restrict linkage to external data sources, impeding integration of comprehensive clinical phenotypes and real-world outcomes. These limitations emphasize the need for diverse, longitudinal, deeply phenotyped cohorts, standardization, and experimental validation to advance understanding of autoimmune disease mechanisms and comorbidities.

## 7 Conclusion

This study advances understanding of autoimmune disease (AD) genetics by integrating functional variant analyses, polygenic risk modeling, and cross-cohort evaluation. Polygenic score (PGS) analyses revealed both shared and divergent genetic architectures across ADs, highlighting partial genetic overlap rather than complete convergence. Positive odds ratios and risk correlations underscore the interconnected nature of autoimmune pathophysiology and the clinical importance of comorbidity. Moderate overlapping susceptibility, rather than extreme risk in one condition, appears to underlie disease co-occurrence. Sex differences remain a defining feature, with a consistent female predominance across most ADs and higher male prevalence in ankylosing spondylitis. These patterns reinforce sex as a critical biological covariate influenced by hormonal, genetic, and diagnostic factors. Differential dependence on the HLA region among diseases further demonstrates the need for disease-specific analytic strategies, some conditions retaining strong non-HLA signals, others dominated by HLA-linked variation. Integrated analyses of common and rare variants identified both established and novel gene disease associations, expanding the genetic landscape of autoimmunity. Network propagation revealed systemic versus local pathway perturbations, offering a functional framework for interpreting genetic risk. Cross-database comparisons between TriNetX and UK Biobank highlight three key challenges: inconsistencies in ICD-10 coding and disease terminology that limit data harmonization and comparability, sex-biased prevalence patterns, and dataset-specific variability in disease-pair associations. These differences underscore the necessity of harmonised diagnostic standards, ancestry-balanced cohorts, and meta-analytic frameworks to ensure reproducibility and accurate interpretation of epidemiological signals. Overall, this work provides a harmonised, data-driven resource that links genetic architecture, comorbidity patterns, and methodological robustness in an ancestry-aware context. It delineates both the shared and distinct features of ADs, thereby providing a foundation for precision medicine approaches that leverage common biological mechanisms while acknowledging disease-specific nuances.

## Data Availability

The data used in this study were obtained from the UK Biobank resource under application number 91246 and from the TriNetX research network. Restrictions apply to the availability of these data, which were used under license/access agreements for the current study and are therefore not publicly available. Researchers may apply for access to UK Biobank and TriNetX data through their respective access procedures. Study-generated summary data and supplementary tables are provided with this manuscript in the accompanying supplementary files

## Funding

HB and RS gratefully acknowledge funding from the Deutsche Forschungsgemeinschaft (DFG, German Research Foundation) under Germany’s Excellence Strategy (EXC 22167-390884018) and by Research Training Group 2633 “Autoimmune Pre-Disease,” project A9 (GRK 2633/9 – 2021). Open Access funding was enabled and organized by Projekt DEAL.

## Notes

### Competing Interest Statement

The authors have declared no competing interest.

### Author Declarations

The North West Multi-centre Research Ethics Committee gave ethical approval for the UK Biobank resource. This study used de-identified data obtained from UK Biobank under approved access procedures. This research was conducted using the UK Biobank resource under application number 91246. TriNetX provided access to de-identified patient data in compliance with HIPAA privacy standards. The use of de-identified data and waiver of informed consent were consistent with applicable ethical and regulatory guidelines.

