## Supplemental File 1 for "Genetic Profiling of Autoimmune Diseases and Exploring Clusters Through Polygenic Risk Score Analysis Using Cohort Data from the UK Biobank"

### 1 Supplementary

#### 1.1 Distributional Details for Genetic Risk Scores in the Enhanced Dataset

In the case of MS and PSO (Figure 3A in the main manuscript) 47% of the samples are male and 53% are female. In the top-left quadrant, MS is the preponderance (46%), while it accounts for 28% in the bottom-right and bottom-left quadrants. PSO occupies the bottom-right quadrant (38%), and the bottom and top-left quadrants collectively contain 38% of PSO samples. MS and PSO are similarly represented in the top-right quadrant, with 26% and 24% of samples, respectively. In quadrants that are dominated by MS, female representation is higher at 75%, whereas the PSO quadrants have a nearly equal gender ratio at 50.4% female. More than 50% of the samples are distributed across the four quadrants in both diseases.

Similarly, 29.8% of the MS and RA samples are male, while 70.2% female (Figure 3B in the main manuscript). MS continues to dominate the top-left quadrant, accounting for 47% of the samples. On the other hand, the bottom-right and bottom-left quadrants have 28%. The bottom-right quadrant has the most RA samples (37%), and the bottom and top-left quadrants together have 39% of RA samples. Both diseases share the top-right quadrant, with 25% of the samples coming from MS and 23% from RA. The high-percentage quadrants for both diseases are primarily female, with 80% of samples in the top-left quadrant dominated by MS and 70% in the bottom-right quadrant dominated by RA belonging to females. In both MS and RA, female representation exceeds male representation in all quadrants, and more than half of the total samples are distributed across various quadrants in each disease comparison.

In MS and SLE comparison (Figure 3C in the main manuscript), 20% of the patients in the sample are male, while 80% are female. The top right quadrant contains the maximum number of samples for both diseases, with 46% of samples for MS and 41% for SLE. In Comparison between PSO and RA (Figure 3D in the main manuscript), The greatest proportion of female samples is RA, with male and female participants comprising 41% and 59%, respectively. The uppermost left and right quadrants contain approximately 62% of the PSO samples. Half of the 62% of samples have high PGS for both diseases; however, the other half have a low-risk score for RA but a high PGS for PSO. 35% of the samples are located in the bottom-right quadrant for RA, with 26% in the top-right quadrant. The top-right quadrant (33%) has the most samples with both disorders, followed closely by the top-left quadrant (27%) and the bottom-right quadrant (24%).

The ADs PSO and SLE (Figure 3E in the main manuscript) analysis included 47% men and 53% women. For PSO, the distribution is somewhat balanced between males and females, with both sexes evenly distributed in all quadrants. The top-left quadrant has the most overall representation (35%), and most of the PSO samples are in line with higher Y-axis values. SLE, on the other hand, has a high gender bias, with 96% of the samples in the top-right quadrant being women. SLE samples are primarily located in the bottom-right quadrant (44%), with a preference for higher X-axis values. While PSO has a more uniform distribution across quadrants, SLE has a more skewed pattern, affected primarily by female samples.

The RA and SLE pair of AD analyses (Figure 3F in the main manuscript) included 29.1% male and 70% female subjects. For RA and SLE, there is a considerable female bias, with 70% in the top-left quadrant and 92% in the bottom-right quadrant. Notably, the top-right quadrant has the highest overall representation: 32% for RA and 33% for SLE. Each disease has a distribution across multiple quadrants and a more polarized pattern, which is heavily influenced by female samples. In comorbid samples for both sexes, the majority of instances (57%) were located in the top-right quadrant. Despite the fact that the comorbidity group had a higher mean than SLE samples but a lower mean than RA samples.

#### 1.2 Detailed Pathway Enrichment Outputs from Network Propagation Analysis

Among the 50 evaluated hallmark pathways, CED shows positive scores for 42 pathways (pathways related to immune and inflammatory signalling, cell-cycle control and proliferative/oncogenic programmes, metabolic and bio-energetic pathways, stress-adaptation and damage-response modules, developmental and lineage-specification pathways, cellular architecture and polarity), the highly positive (more than 0.50) pathways are related to Proliferation, Immune and Development and negative scores for 7 pathways (core energy metabolism, prosthetic-group/iron handling, oxygen-sensing response, lineage differentiation, secretory machinery, oxidative-stress defence). CD records 32 positive and 18 negative pathways, positive assignments include pathway related to hormone responses, metabolism, immune and inflammatory signaling, growth-factor and oncogenic signaling, Genome Integrity and Stress, Differentiation and Morphogenesis; all remaining pathways are negative mostally related to cell architecture and polarity, cell cycle and proliferation, signal transduction. PSO exhibits positive scores for 9 pathways related to cell cycle and proliferation, genome integrity and stress, secretory pathways, differentiation and morphogenesis and negative for 35 pathways mostly related to immune and inflammatory, metabolism and detoxification, signaling and development, cell cycle and stress response. RA displays 22 positive pathways related to signaling and development, immune and inflammation, tissue structure and function, cell cycle, stress and metabolism and 25 negative pathways related to cell cycle and genome integrity, metabolism and homeostasis, signaling and stress response, differentiation and tissue function. Similarly, SLE has positive status for 39 pathways, highest positive (more than 0.55) are metabolic pathway. SLE has negative status for 11, which includes signaling and development, cell cycle and proliferation, stress and quality control, secretion and specialized function. T1D yields 42 positive pathways related to metabolism and homeostasis,immune and vascular, signaling and stress, structure and differentiation, there are 8 negative pathways mainly related to cell cycle and proliferation, genome integrity and repair, proteostasis and stress response, differentiation and tissue function. UC registers positive for 31 pathways immune and inflammation, metabolism and homeostasis, cell cycle and stress response, development and differentiation and negative for 18 pathways includeing signaling, development, cell cycle, proliferation, metabolism, detoxification, structure, secretion and stress response.

Genes related to CED is positive for 13 pathways allograft rejection, androgen response, angiogenesis, apical junction, apoptosis, bile acid metabolism, cholesterol homeostasis, coagulation, IL6 jak stat3 signalling, inflammatory response, interferon gamma response, TGF-beta signalling and the reactive oxygen species pathway. CED genes are negatively enriched for adipogenesis and oxidative phosphorylation. CD shows 10 positive assignments in adipogenesis, allograft rejection, angiogenesis, apoptosis, bile acid metabolism, IL6 jak stat3 signalling, inflammatory response, interferon gamma response, TGF-beta signalling and oxidative phosphorylation. There are 5 negatively enriched pathways (androgen response, apical junction, cholesterol homeostasis, coagulation, reactive oxygen species) are related to genes identified for CD. PSO records two high negative values (angiogenesis, IL6 jak stat3 signalling) and negative status for the remaining 13 pathways, with no positive enrichment. RA has 5 positively enriched pathways (allograft rejection, apical junction, inflammatory response, interferon gamma response, TGF-beta signalling) and 10 negatively enriched pathways. Systemic lupus erythematosus is positive for 14 pathways (all except TGF-beta signalling) and negative for that single pathway. T1D is positive for 14 pathways, including one high-positive (IL-6 jak stat3 signalling), with no negative enrichemt. UC registers 10 positive pathways (adipogenesis, allograft rejection, angiogenesis, apoptosis, bile acid metabolism, IL-6 jak stat3 signalling, inflammatory response, interferon gamma response, TGF-beta signalling, oxidative phosphorylation) and 5 negatively enriched pathways (androgen response, apical junction, cholesterol homeostasis, coagulation, reactive oxygen species).

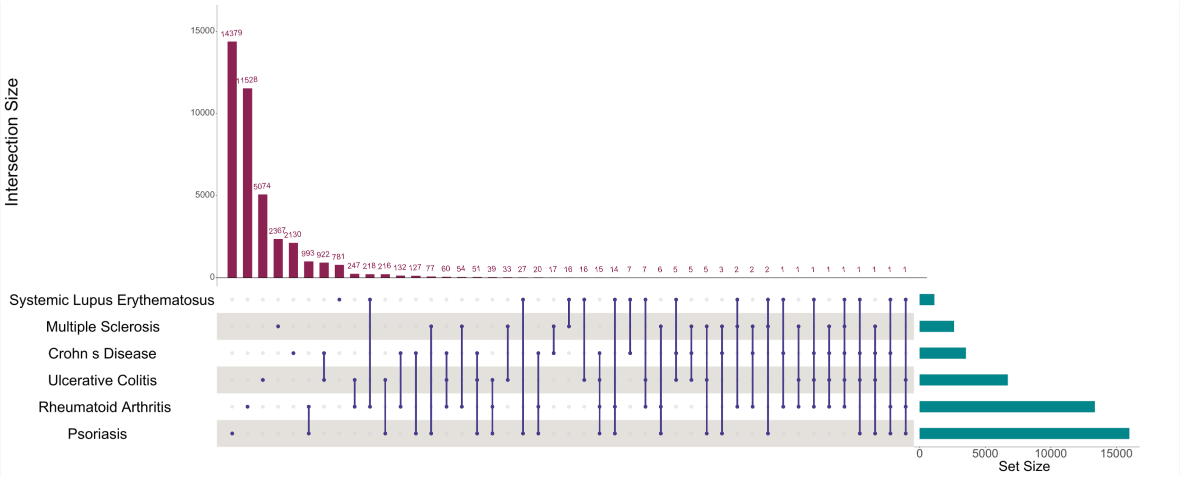


**Figure S1:** Upset plot illustrates the overlap of samples among ADs using first reported (phenotype) data from the UKB. The dark red bar chart quantifies each intersection, with the numbers top each bar indicating the sample count shared between the sets. Each row represents an original set, while each column corresponds to a specific intersection. Within the matrix, blue filled dots denote the presence of a set in a given intersection, and empty spaces indicate its absence. Additionally, a separate dark green section displays the total size of each individual set for reference.

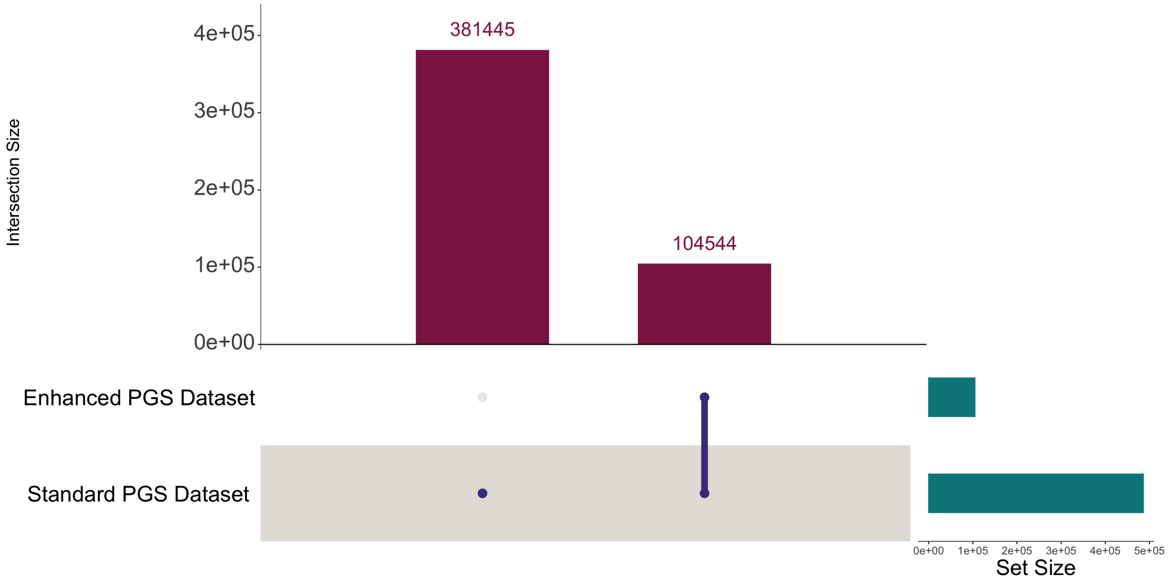


**Figure S2:** An Upset plot representing the number of overlap samples between standard and enhanced dataset. The bar chart (dark red) shows the size of each intersection indicating how many samples are shared among the two sets. Each row represents one of the original sets and each column represents a specific intersection. In the matrix, filled dots (blue) indicate the sets that contribute to that particular intersection, while empty spaces denote the absence of a set in that combination. Additionally, the separate section (dark green) displays the overall size of each individual set for reference. The plot illustrates that the enhanced dataset is a subset of the standard dataset.

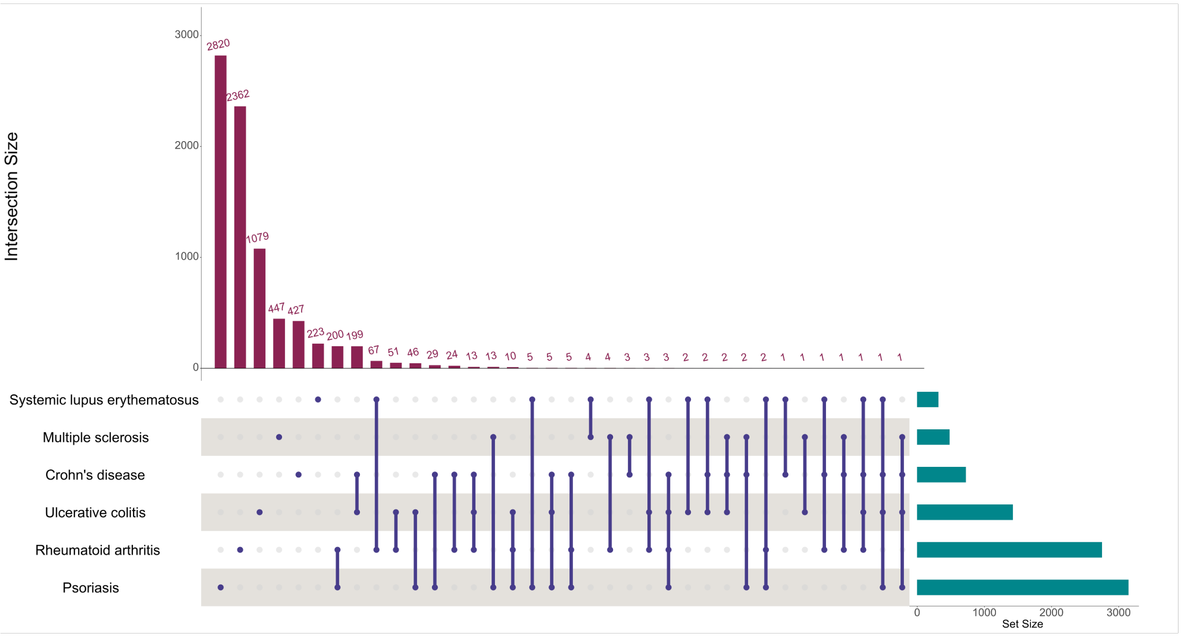


**Figure S3:** This upset plot visualizes the overlap of samples among ADs in the enhanced dataset. The dark red bar chart represents the size of each intersection, with the numbers above the bars indicating the sample count shared between sets. Each row corresponds to an original set, while each column represents a specific intersection. In the matrix, blue filled dots signify the presence of a set within an intersection, whereas empty spaces indicate its absence. Additionally, a separate dark green section highlights the total size of each individual set for reference.

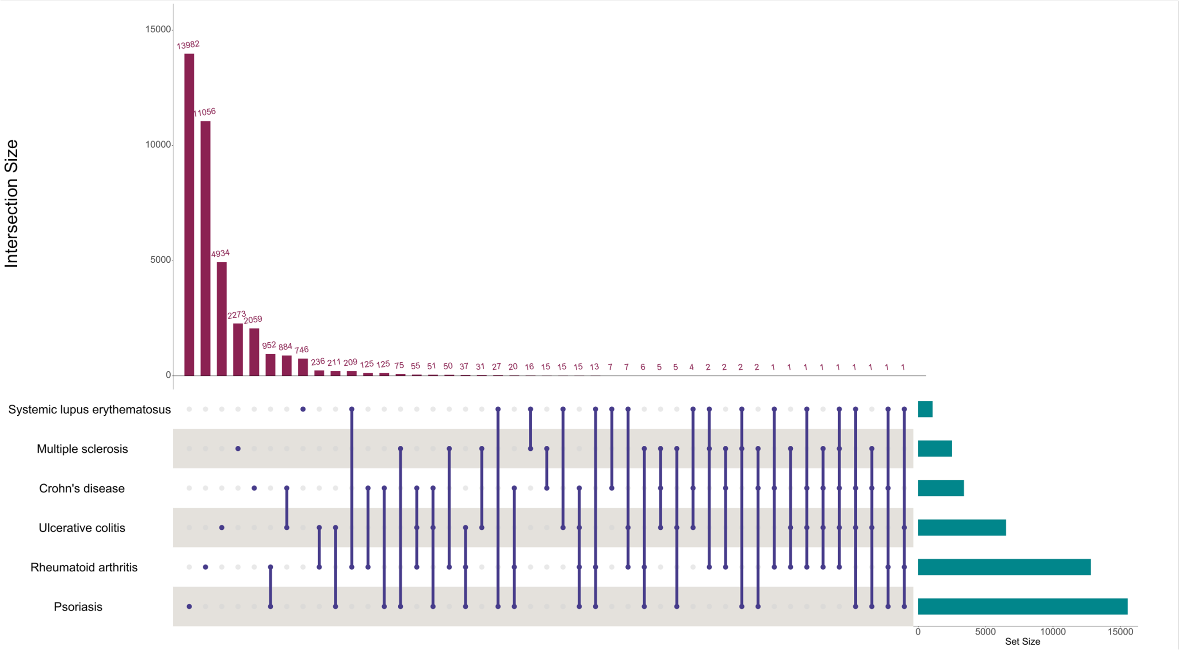


**Figure S4:** This upset plot visualizes the overlap of samples among ADs in the standard dataset. The dark red bar chart represents the size of each intersection, with the numbers above the bars indicating the sample count shared between sets. Each row corresponds to an original set, while each column represents a specific intersection. In the matrix, blue filled dots signify the presence of a set within an intersection, whereas empty spaces indicate its absence. Additionally, a separate dark green section highlights the total size of each individual set for reference.

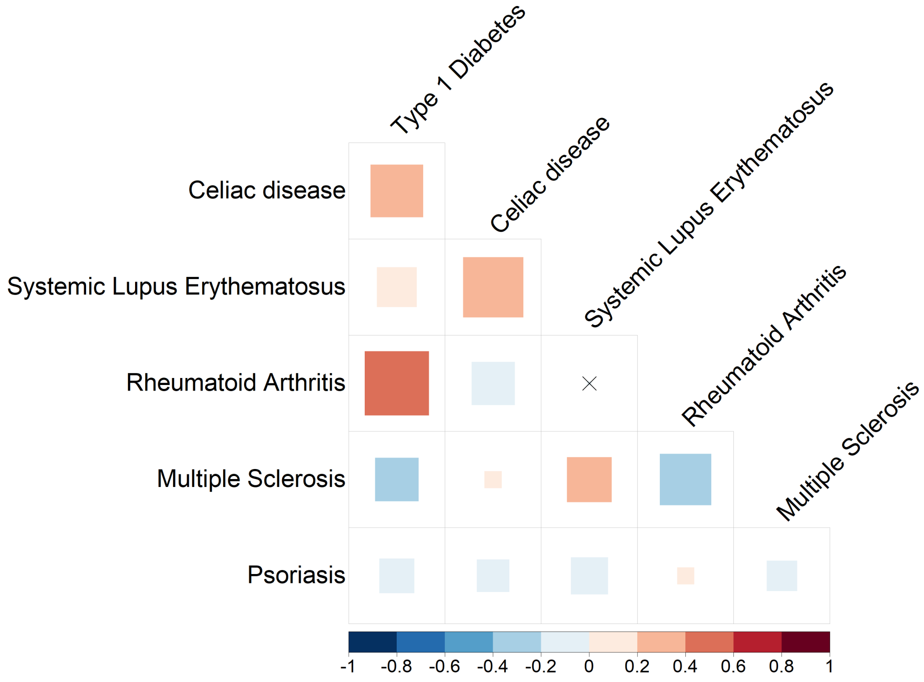


**Figure S5:** Correlation plot to visualize the strength and direction of relationships between various traits, diseases, and ADs. The analysis utilises PGS derived from the Enhanced dataset. Brown denotes traits, green presents diseases, and pink denotes autoimmune conditions. The plot employs a colour gradient to indicate correlation strength, with red representing positive correlations (more than 0) and blue indicating negative correlations (less than 0). No association is indicated by lighter (near 0 and white) colours. The correlation coefficient, ranging from -1 to 1, quantifies the relationship, with values closer to 1 suggesting a strong positive correlation and values near -1 indicating a strong negative correlation. Each cell in the matrix corresponds to a pairwise comparison, facilitating the identification of highly correlated clusters of variables. The cross mark denotes correlations that are not statistically significant.

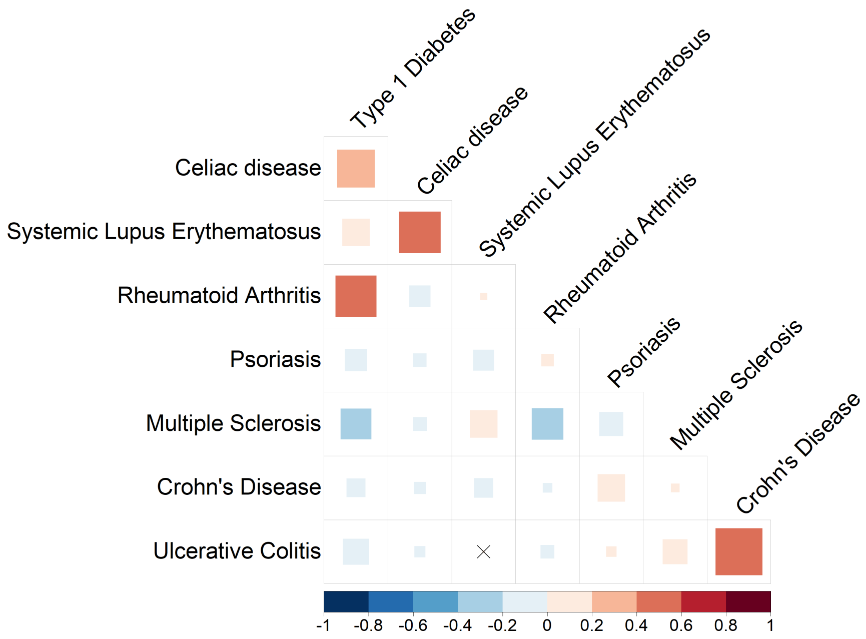


**Figure S6:** Correlation plot visualizes the strength and direction of associations between traits, diseases, and autoimmune conditions using PGS calculated from the Standard dataset. Variables are color-coded by category: traits (brown), diseases (green), and ADs (pink). Correlation strength is depicted through a gradient, with red signalling positive relationships (more than 0), blue indicating negative relationships (less than 0), and lighter shades (near 0 and white) representing negligible associations. Numerical correlation coefficients (ranging from -1 to 1) quantify these relationships, where values approaching 1 or -1 denote robust positive or negative links, respectively. Each cell in the matrix corresponds to a pairwise comparison, highlighting clusters of interconnected variables. Statistically non-significant correlations are marked with cross symbols to distinguish them from meaningful associations.

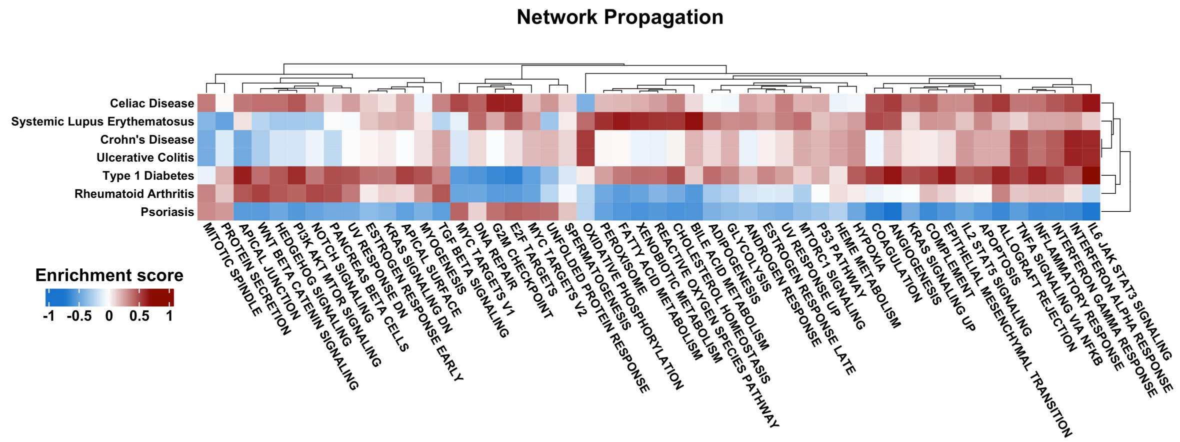


**Figure S7:** The heatmap depicts the results of network propagation, highlighting the associations between biological pathways and diseases based on genes selected through rare, common and impact based filtering. In the plot, rows represent pathways and columns correspond to diseases. Hierarchical clustering is applied, with dendrograms along the top and left to illustrate relationships among pathways and diseases. The color gradient indicates the magnitude of variant effects on each pathway, with enrichment scores ranging from −1 to 1. Pathways with negative enrichment are less influenced by associated variants, whereas those with positive enrichment are more likely to contribute significantly to disease mechanisms.

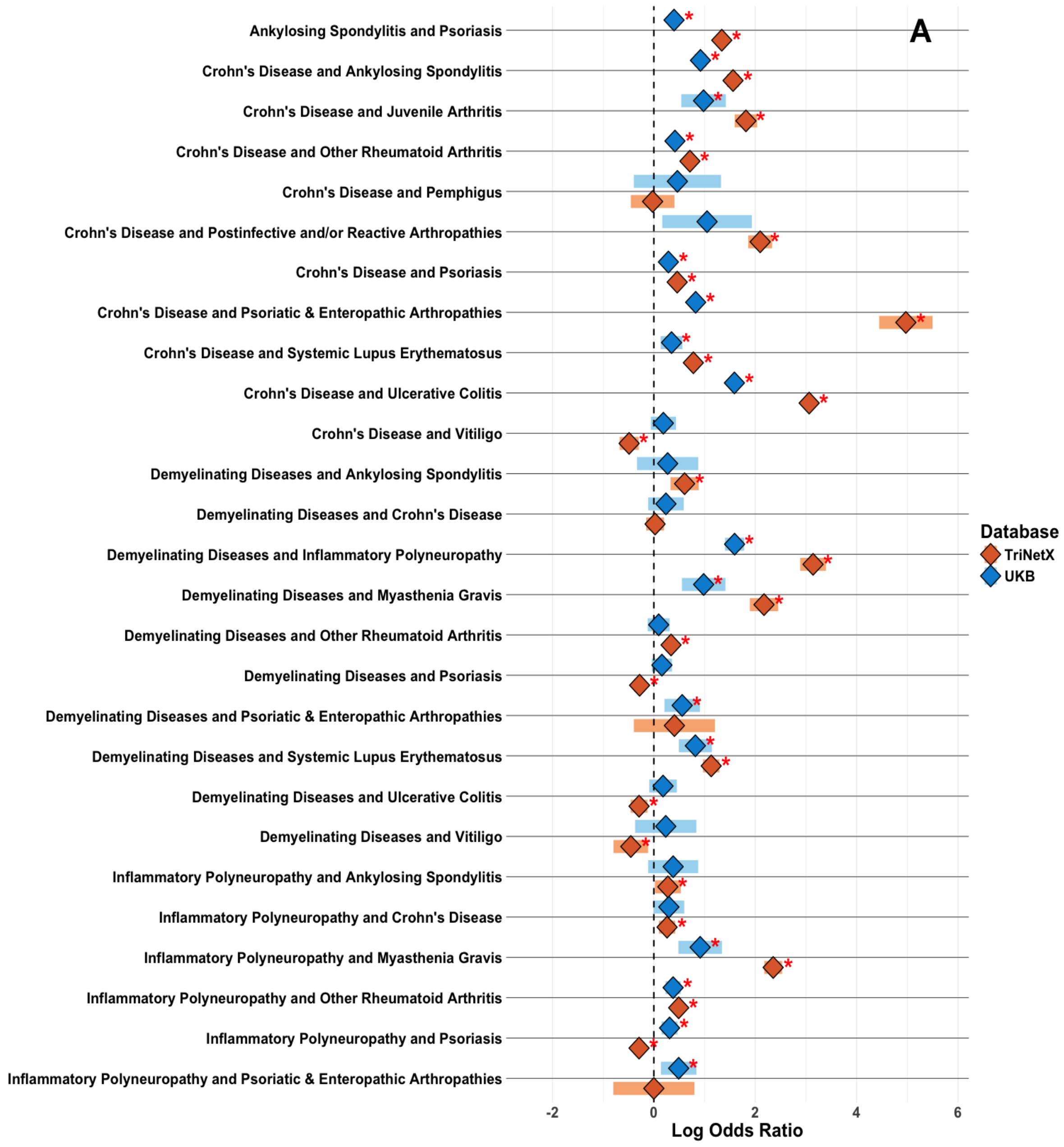
A
Ankylosing Spondylitis and Psoriasis
Crohn's Disease and Ankylosing Spondylitis
Crohn's Disease and Juvenile Arthritis
Crohn's Disease and Other Rheumatoid Arthritis
Crohn's Disease and Pemphigus
Crohn's Disease and Postinfective and/or Reactive Arthropathies
Crohn's Disease and Psoriasis
Crohn's Disease and Psoriatic & Enteropathic Arthropathies
Crohn's Disease and Systemic Lupus Erythematosus
Crohn's Disease and Ulcerative Colitis
Crohn's Disease and Vitiligo
Demyelinating Diseases and Ankylosing Spondylitis
Demyelinating Diseases and Crohn's Disease
Demyelinating Diseases and Inflammatory Polyneuropathy
Demyelinating Diseases and Myasthenia Gravis
Demyelinating Diseases and Other Rheumatoid Arthritis
Demyelinating Diseases and Psoriasis
Demyelinating Diseases and Psoriatic & Enteropathic Arthropathies
Demyelinating Diseases and Systemic Lupus Erythematosus
Demyelinating Diseases and Ulcerative Colitis
Demyelinating Diseases and Vitiligo
Inflammatory Polyneuropathy and Ankylosing Spondylitis
Inflammatory Polyneuropathy and Crohn's Disease
Inflammatory Polyneuropathy and Myasthenia Gravis
Inflammatory Polyneuropathy and Other Rheumatoid Arthritis
Inflammatory Polyneuropathy and Psoriasis
Inflammatory Polyneuropathy and Psoriatic & Enteropathic Arthropathies
-2
0
2
4
6
Log Odds Ratio
Database
TriNetX
UKB

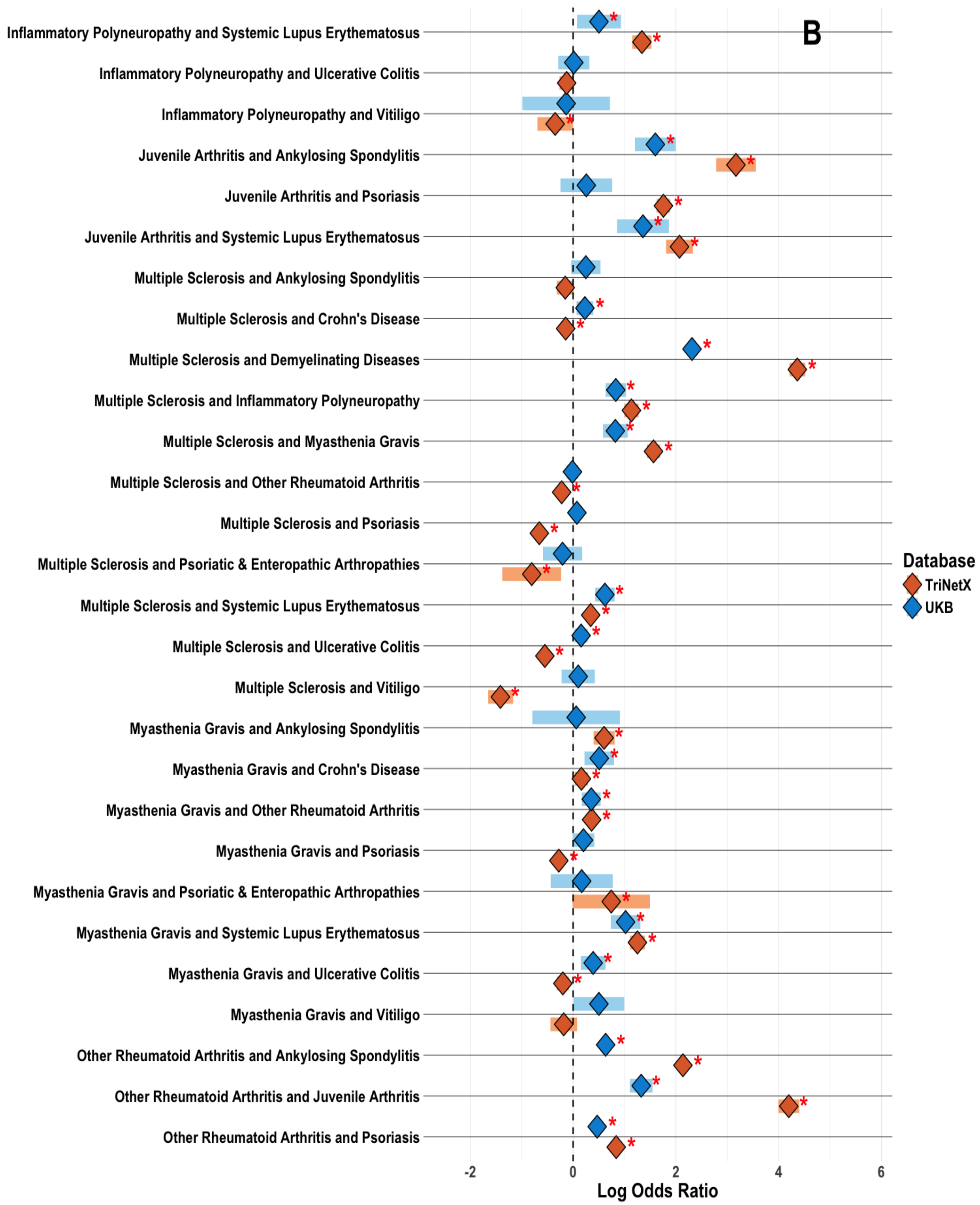
B
Inflammatory Polyneuropathy and Systemic Lupus Erythematosus
Inflammatory Polyneuropathy and Ulcerative Colitis
Inflammatory Polyneuropathy and Vitiligo
Juvenile Arthritis and Ankylosing Spondylitis
Juvenile Arthritis and Psoriasis
Juvenile Arthritis and Systemic Lupus Erythematosus
Multiple Sclerosis and Ankylosing Spondylitis
Multiple Sclerosis and Crohn's Disease
Multiple Sclerosis and Demyelinating Diseases
Multiple Sclerosis and Inflammatory Polyneuropathy
Multiple Sclerosis and Myasthenia Gravis
Multiple Sclerosis and Other Rheumatoid Arthritis
Multiple Sclerosis and Psoriasis
Multiple Sclerosis and Psoriatic & Enteropathic Arthropathies
Multiple Sclerosis and Systemic Lupus Erythematosus
Multiple Sclerosis and Ulcerative Colitis
Multiple Sclerosis and Vitiligo
Myasthenia Gravis and Ankylosing Spondylitis
Myasthenia Gravis and Crohn's Disease
Myasthenia Gravis and Other Rheumatoid Arthritis
Myasthenia Gravis and Psoriasis
Myasthenia Gravis and Psoriatic & Enteropathic Arthropathies
Myasthenia Gravis and Systemic Lupus Erythematosus
Myasthenia Gravis and Ulcerative Colitis
Myasthenia Gravis and Vitiligo
Other Rheumatoid Arthritis and Ankylosing Spondylitis
Other Rheumatoid Arthritis and Juvenile Arthritis
Other Rheumatoid Arthritis and Psoriasis
-2
0
2
4
6
Log Odds Ratio
Database
TriNetX
UKB

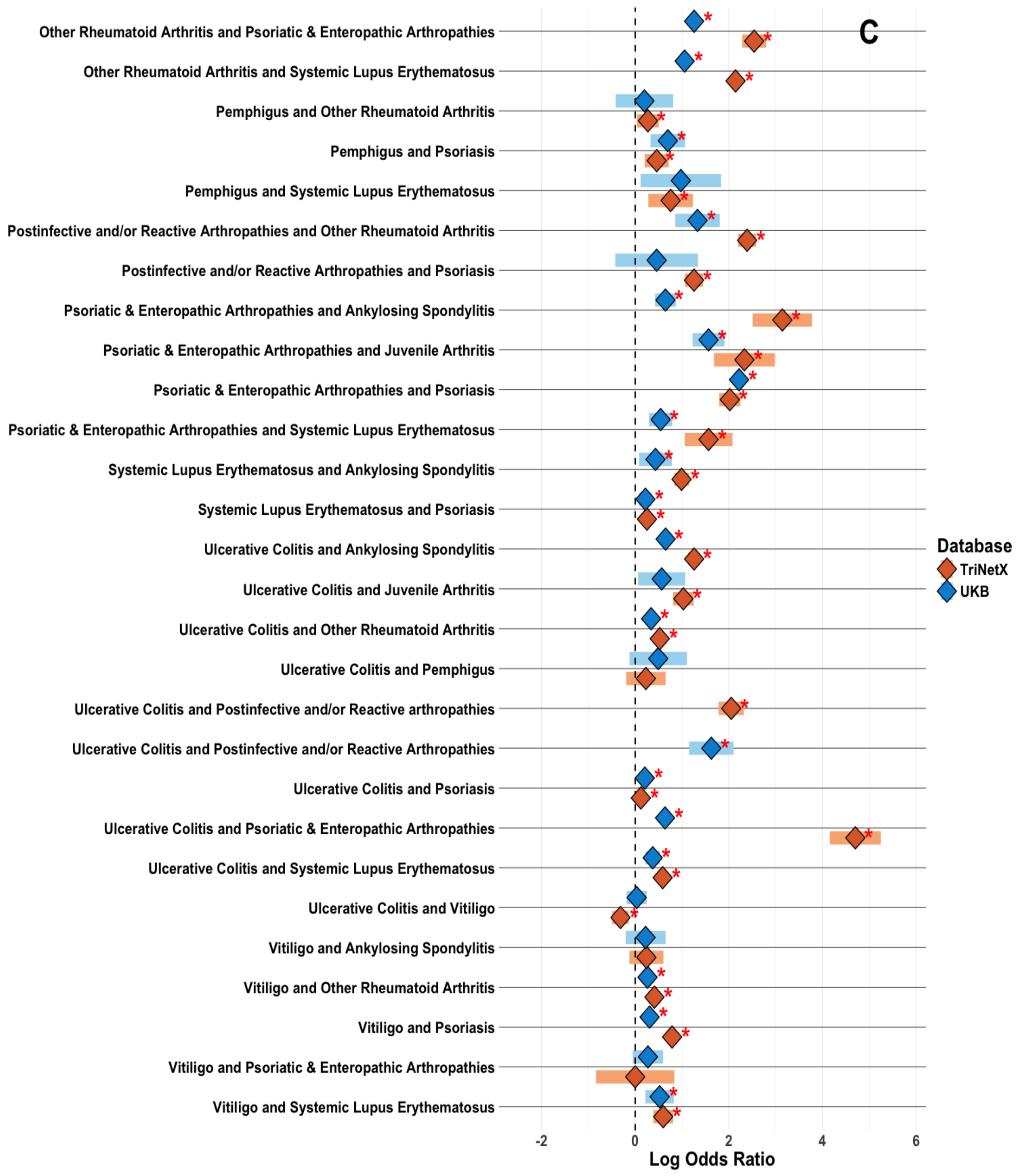


**Figure S8:** Combined figure A, B and C presents log odds ratios (OR) for combinations of diseases, ranging from -2 to 6. Orange bars denote odds ratios obtained from TNX, while blue bars represent those from the UKB. A star symbol denotes statistically significant OR, and the lines surrounding each OR indicate the corresponding confidence intervals
